# Estrogen-related receptor genes underlie sex differences in cortical atrophy associated with isolated REM sleep behavior disorder

**DOI:** 10.1101/2025.04.14.25325684

**Authors:** Marie Filiatrault, Violette Ayral, Christina Tremblay, Celine Haddad, Véronique Daneault, Alexandre Pastor-Bernier, Jean-François Gagnon, Ronald B. Postuma, Petr Dusek, Stanislav Marecek, Zsoka Varga, Johannes Klein, Michele T. Hu, Isabelle Arnulf, Pauline Dodet, Marie Vidailhet, Jean-Christophe Corvol, Stéphane Lehéricy, ICEBERG Study Group, Simon Lewis, Elie Matar, Kaylena A. Ehgoetz Martens, Lachlan Churchill, Per Borghammer, Karoline Knudsen, Allan K. Hansen, Dario Arnaldi, Beatrice Orso, Pietro Mattioli, Luca Roccatagliata, Shady Rahayel, Quebec Parkinson Network

## Abstract

Isolated REM sleep behavior (iRBD) is a male predominant parasomnia characterized by abnormal dream-enacting movements in REM sleep. It is the prodromal manifestation most strongly associated with the development of synucleinopathies, such as Parkinson’s disease or dementia with Lewy bodies. While individuals with iRBD exhibit significant cortical atrophy shaped by distinct gene expression, sex-specific differences in structural brain changes remain unknown. In this study, we investigate the effect of sex on brain atrophy in iRBD and examine the gene expression underpinning the brain abnormalities in a large international multicentric MRI dataset with polysomnography-confirmed iRBD.

T1-weighted scans from 408 individuals with iRBD and 480 healthy controls were acquired. Vertex-based cortical surface reconstruction and segmentation were conducted, and general linear models were used to quantify brain atrophy and assess the sex effect on cortical thickness in iRBD compared to controls. We then used a high resolution parcellation to further characterize the sex differences and conduct imaging transcriptomics analyses. Gene enrichment analyses were performed to identify genes associated with sex differences in cortical atrophy in iRBD.

Males with iRBD showed significantly more cortical thinning compared to females with iRBD and controls, despite similar age and clinical features. The gene enrichment analysis revealed that female-specific resilience in cortical atrophy was associated with overexpression of oestrogen-related receptors.

These findings provide mechanistic insight of sex-specific neuroprotection in prodromal stages of synucleinopathies, highlighting the critical impact of sex on the progression of neurodegenerative diseases.

## Introduction

Isolated REM sleep behavior disorder (iRBD) is a parasomnia characterized by the loss of REM sleep muscle atonia, leading to dream-enacting behaviors such as vocalizations, limb movements, and complex, often violent, movements that closely align with dream content.^1,2^ Critically, iRBD is the strongest known prodromal marker of neurodegenerative synucleinopathies, with over 90% of individuals eventually developing dementia with Lewy bodies (DLB), Parkinson’s disease (PD), or in a smaller proportion, multiple system atrophy (MSA).^3^ Phenoconversion can take up to 15 years,^3,4^ offering an important window for studying early biomarkers and disease mechanisms before overt neurodegeneration emerges.^5^

Neuroimaging studies show that individuals with iRBD already exhibit cortical atrophy compared to healthy controls,^6–8^ which are associated with cognitive decline and motor impairments.^9,10^ While definite phenoconversion biomarkers are still being investigated, a specific atrophy signature can predict whether an individual with iRBD is more likely to convert to DLB rather than PD.^11^ Brain atrophy, such as reduction in cortical thickness, in iRBD follows distinct patterns, potentially driven by prion-like propagation along neural networks and selective regional vulnerability,^8^ with the most affected areas overexpressing genes related to mitochondrial function and macroautophagy.^12^

Sex is another key factor influencing neurodegeneration,^13^ yet its role in brain changes associated with iRBD remains largely unexplored. iRBD has a strong male predominance, with reported prevalence ratios as high as 8:1,^4,14^ leading to an underrepresentation of female individuals in iRBD studies, particularly in neuroimaging studies. In manifest synucleinopathies, sex differences are well documented.^15^ In DLB and PD,^16,17^ studies highlight that males show earlier onset,^17,18^ greater motor impairment,^18,19^ and more extensive and severe neurodegenerative brain changes,^20–22^ while females exhibit relative neuroprotection, possibly mediated by oestrogens.^13,23^ Estrogens and estrogen-related functions play a key role in mitochondrial function and dopaminergic neuron survival,^24,25^ potentially delaying neurodegeneration. These protective effects, supported by human and animal studies,^26,27^ have been suggested as a possible explanation for the milder cortical atrophy, slower and more benign disease progression in females compared to males with DLB and PD.^13,28^ However, whether such sex-related neuroprotective mechanisms already operate in early synucleinopathies remains unknown.

Studies on sex effects in iRBD are scarce and yield conflicting results.^29^ While some studies suggest that females have a later onset age,^30^ others report earlier onset^31^ or no difference.^14^ Some behavioural studies indicate that females exhibit less aggressive dream-enacting behaviours and fewer sleep-related injuries compared to males,^31–33^ though findings are inconsistent.^15^ Sleep architecture differences such as shorter N1 sleep and longer REM latency in females have also been noted.^30,34^ However, beyond these observations, large longitudinal studies have failed to detect sex-related differences in disease progression,^35,4^ leaving a gap in understanding how sex influences disease trajectory. Neuroimaging findings suggest a more complex interplay between sex and brain changes in iRBD. Our group previously identified atrophy and perfusion signatures predictive of phenoconversion in iRBD.^13,37^ While the atrophy signature predicting DLB did not show sex differences, deformation in brainstem tissue associated with REM sleep motor activity was more prominent in males.^11^ Moreover, a signature of perfusion changes predicting phenoconversion risk in iRBD was also more pronounced in males.^36^ These findings highlight the need for further research into sex-specific neurodegenerative patterns in iRBD.

In this study, we used a large international, multicentre dataset of 888 brain MRI scans (408 polysomnography-confirmed iRBD patients and 480 healthy controls) to investigate sex-related differences in atrophy. Using vertex-based cortical surface analysis, we investigated the presence of a sex interaction on cortical thickness between iRBD patients and controls. We quantified the extent of cortical thinning in male and female iRBD patients matched for age and clinical features and used imaging transcriptomics to investigate whether the spatial distribution of gene expression in the brain underlies sex-related atrophy differences. Gene enrichment analysis was further performed to identify relevant molecular pathways in regions with sex-related atrophy effects in iRBD. To further investigate the systemic relevance of genes implicated in sex-related cortical atrophy, we examined their normal expression patterns across peripheral and central tissues from the Genotype-Tissue Expression (GTEx) Project. We hypothesized that males with iRBD would exhibit greater cortical thinning than females, with regions less affected in females overexpressing genes involved in estrogen-related molecular functions.

## Methods

### Participants

A total of 888 participants were recruited for this study and underwent T1-weighted brain MRI imaging. This cohort included 408 polysomnography-confirmed iRBD patients and 480 age-matched healthy controls, recruited from nine international centers: 179 (85 patients) from the Centre for Advanced Research on Sleep Medicine at the Hôpital du Sacré-Cœur de Montréal, Montreal, Canada; 140 (83 patients) from the First Faculty of Medicine at Charles University, Prague, Czechia; 147 (81 patients) from the Oxford Discovery Cohort, Oxford, UK; 136 (60 patients) from the Movement Disorders Clinic (ICEBERG and ALICE cohorts) at the Hôpital de la Pitié-Salpêtrière, Paris, France; 56 (30 patients) from the ForeFront Parkinson’s Disease Research Clinic, Sydney, Australia; 38 (18 patients) from Aarhus University Hospital, Aarhus, Denmark; 29 (14 patients) from IRCCS Ospedale Policlinico San Martino, Genoa, Italy; and 163 (37 patients) from the Parkinson’s Progression Markers Initiative (PPMI) baseline cohort.^37^ These participants were previously included in a machine learning study investigating brain atrophy progression pathways in synucleinopathies.^38^

All iRBD patients underwent video-polysomnography and were diagnosed based on the International Classification of Sleep Disorders criteria.^2^ Neurological evaluations and cognitive assessments confirmed that patients were still in the isolated phase of RBD. Patients were excluded if they had, at the clinical visit closest in time to the MRI session, a diagnosis of DLB, PD or MSA based on published diagnostic criteria,^39–41^ had a history of brainstem stroke, epilepsy or epileptiform activity on EEG, had antidepressant-triggered RBD, had untreated obstructive sleep apnea, or had RBD mimics such as sleepwalking and night terrors. All patients underwent standardized clinical evaluations, including motor assessments using the Movement Disorders Society-sponsored Unified Parkinson’s Disease Rating Scale (MDS-UPDRS-III)^42^ and global cognitive evaluation using the Montreal Cognitive Assessment (MoCA).^43^All patients were part of ongoing prospective studies. Study protocols were approved by the Research Ethics Board of the Quebec Integrated University Centre for Health and Social Services of Northern Island of Montreal, the McGill University Health Centre, and the respective local ethics boards at all participating sites.

### MRI acquisition and processing

T1-weighted MRI scans were acquired using 3T MRI scanners across the different sites, with site-specific acquisition protocols (detailed in the Supplementary Material). To assess sex-related effects on cortical morphology in iRBD, MRI scans were processed to generate whole-brain vertex-based cortical thickness maps using FreeSurfer (version 7.1.1). The standard processing pipeline included intensity normalization, brain extraction, segmentation of subcortical structures, cortical surface reconstruction, and topological correction (technical details in Supplementary Material). Cortical thickness was calculated as the closest distance between the grey-white and grey-CSF boundaries at each vertex, producing high-resolution maps sensitive to submillimeter cortical differences. All cortical reconstructions were visually inspected by trained raters (S.R. and V.A.) and scored based on established guidelines,^44,45^ as done previously.^8,12^ Secondary analyses were conducted on cortical surface area and cortical volume maps, with surface area defined as the sum of triangle areas at each vertex and cortical volume as the product of cortical thickness and surface area.

To characterize brain regions showing significant sex effects on cortical thickness and performing gene expression analysis, cortical maps were parcellated. Unlike previous studies that used the standard 34-region Desikan-Killiany atlas,^46^ we applied the Lausanne atlas with a higher-resolution, including 1000 cortical regions, to improve spatial granularity of sex effects on atrophy.^47^ The fetch_cammoun2012 function from netneurotools^47^ was used to retrieve atlas annotation files in fsaverage5 template space, which were then registered to each participant’s cortical surface via FreeSurfer’s mri_surf2surf function. The aligned annotation files were converted to gifti format using mris_convert for compatibility with downstream processing, and cortical thickness measures were extracted for each parcellated region with custom scripts. To account for scanner-specific effects in this multicentric dataset, cortical thickness values were harmonized across acquisition sites using the ComBat tool,^48^ an empirical Bayes-based approach originally developed for genomics^49^ and widely applied in multicentric MRI studies, including our previous work.^8,12,50^ ComBat correction was applied while preserving biological variability of interest (i.e., group, age, and sex), thereby removing variance attributable to site-specific effects. To ensure that age effects did not confound the atrophy-gene expression analyses, we applied W-scoring, a normalization technique used to remove age and sex effects in normal aging.^8,12,21^ For each iRBD patient, W-scores were computed for all parcellated regions using linear regression models with the healthy control data, adjusting for age and sex. W-scores represent the standardized difference between observed and expected cortical thickness in iRBD patients, normalized by residual variance.^21^ Negative W-scores indicate greater cortical thinning relative to age– and sex-matched controls. To quantify sex-related effects, we performed an additional set of linear regression analyses on the cortical thickness W-scores across parcellated regions. For each region, the ComBat-corrected W-scores were used as the dependent variable, with sex as the independent variable, producing a single beta coefficient per region. A positive beta indicated that iRBD females exhibited less cortical thinning than iRBD males, whereas a negative beta indicated greater cortical thinning in females. These sex interaction estimates were subsequently used as input in the imaging transcriptomics analysis.

### Gene expression extraction from postmortem brains

To investigate the gene expression patterns underlying sex-related cortical atrophy in iRBD, we extracted gene expression data from the Allen Human Brain Atlas (AHBA) in 1,000 cortical regions (499 regions in the left hemisphere).^47,51^ The AHBA provides gene expression data for over 20,000 genes, quantified across 3,200 tissue samples from six post-mortem healthy adult brains. Gene expression microarray data was accessed using abagen (version 0.1.3), following recommendations for preprocessing and normalization.^52^ Probe-to-gene annotations were verified for accuracy, and probes failing to exceed background noise in at least 50% of samples across donors were discarded.^53^ For genes associated with multiple probes, the probe showing the most stable expression across brain regions was selected. Tissue samples were mapped to the parcellated Lausanne atlas used for atrophy quantification by aligning MNI coordinates, ensuring hemisphere– and structure-specific correspondence (e.g., cortex vs. subcortex). Samples that could not be accurately assigned to a specific brain region were excluded. To mitigate variability across donors, gene expression values were normalized across samples, and regional expression values were averaged first within donors and then across the six donor brains, producing a regional gene expression matrix. Genes with inconsistent expression across donors were excluded. Since the AHBA includes right hemisphere samples from only two donors, previous studies have shown minimal lateralization in microarray expression patterns.^51,54^ Thus, for exploratory reasons, left hemisphere data from the remaining four donors was mirrored to the right hemisphere, though our primary analyses focused on the left hemisphere, as done previously.^12,55^

### Imaging transcriptomics

We applied partial least squares (PLS) regression^56^ to determine whether regional gene expression patterns were associated with sex effects on cortical atrophy in iRBD. PLS regression enabled the identification of latent variables maximizing covariance between the regional cortical thickness sex interaction estimates (for the 499 regions in the left hemisphere) and gene expression levels (15,633 genes across the same 499 regions). Given the high spatial autocorrelation inherent in the brain,^57^ we ensured that gene-atrophy associations were not driven by lower-order spatial gradients by comparing the empirical variance explained by each latent variable to 10,000 spatially constrained null models. Brain regions were randomly shuffled using a spherical reassignment procedure that preserved spatial autocorrelation.^58^ A latent variable was considered significant if fewer than 5% of null models explained more variance than the original atrophy vector. To identify the genes most strongly associated with significant latent variables, we applied a bootstrapping resampling procedure. Rows of the gene expression and cortical thickness matrices were randomly shuffled, and PLS regression was repeated on these shuffled matrices. This procedure was iterated 5,000 times, generating a null distribution and standard errors for each gene’s weight. Bootstrap ratios (the ratio of each gene’s weight to its bootstrap-estimated standard error) were interpreted as *z*-scores,^59^ allowing genes to be ranked from highest to lowest based on their bootstrap ratios. These ranked gene lists were subsequently used as inputs for gene set enrichment analysis.

### Gene set enrichment analysis

To identify enriched functional components in the genes associated with sex-specific cortical atrophy in iRBD, we performed gene set enrichment analysis using WebGestalt 2024.^60^ This analysis aimed to uncover molecular functions, biological processes, and cellular components associated with the brain regions showing sex effects on cortical thinning. Gene terms containing a minimum of 5 and a maximum of 2,000 genes were included in the enrichment analysis. To correct for multiple comparisons, 1,000 random permutations were conducted, with P-values adjusted using the false discovery rate (FDR) method. Significant non-redundant terms associated with less atrophy in iRBD females compared to males (relative to age-matched controls) were interpreted. This approach has demonstrated its efficacy and specificity in neurodegenerative diseases, with atrophy-gene correlations in synucleinopathies highlighting mitochondrial and macroautophagy functions,^12^ protein modelling complexes and *APOE* in Alzheimer’s disease,^12^ and oligodendrocytic cell types in MSA.^61^ To ensure that identified gene terms were not biased by the choice of a specific gene ontology platform, we performed a complementary enrichment analysis using GOrilla^62^ to validate the functional relevance of top-ranked genes.

### Brain vs. peripheral gene expression

To explore whether genes implicated in sex-related cortical atrophy in iRBD were selectively expressed in the central nervous system (specific to brain neurodegeneration) or extended to peripheral tissues, we analyzed gene expression patterns using data from the GTEx Project.^63^ GTEx provides a comprehensive resource for studying human gene expression across 54 non-diseased tissue types, collected from nearly 1,000 post-mortem donors, enabling a detailed assessment of gene expression distribution throughout the body. Gene expression levels for each tissue were obtained from the GTEx Analysis Release V10 (dbGAP Accession phs000424.v10.p2) via the GTEx Portal (accessed 11/22/2024). Expression was quantified using bulk RNA sequencing, reported as transcripts per million (TPM), with isoforms collapsed into a single gene model. For each gene, tissues were classified as overexpressing the gene if their expression exceeded the median expression value across all tissues. To assess whether these genes were disproportionately expressed in the brain compared to the rest of the body, we performed chi-squared tests for each gene, comparing the proportion of overexpressing brain tissues to all other tissue types. Only tissues with available gene expression data for all analyzed genes were included, resulting in 54 distinct tissue types examined.

### Statistical analysis

Group differences in continuous demographic and clinical variables were assessed using independent sample t-tests, while categorical variables were compared using chi-squared tests. Vertex-based cortical thickness analyses were conducted in FreeSurfer using general linear models to assess sex-by-group interactions on cortical thickness, surface area, and volume, adjusting for age and acquisition site. For surface area and cortical volume analyses, estimated total intracranial volume was included as an additional covariate. Analyses were performed separately for the left and right hemispheres, with P-values adjusted for both hemispheres using the 2spaces flag. Surface maps were smoothed with a 15-mm full-width half maximum kernel. Additional vertex-based general linear models were performed to examine sex differences within each group separately. Cluster-level significance was determined using Monte Carlo spatial permutations, with a statistical threshold of P < 0.05 and vertex-level significance set at P < 0.05. To validate our results with a parcel-wise approach, we extracted region-based cortical thickness values from the 1000 cortical region resolution Lausanne parcellation across hemispheres. One-sample t-tests were conducted to compare ComBat-corrected W-scored cortical thickness values of male and female iRBD patients to 0, representing the mean cortical thickness in healthy controls. FDR correction was applied to control for multiple comparisons.^64^ All statistical analyses were performed using SPSS, R, Python, and MATLAB.

## Results

### Demographics and clinical characteristics

Of the 888 participants, 201 (23%) did not pass the quality control, resulting in 687 participants for analysis: 343 iRBD patients and 344 healthy controls (Table 1). The iRBD group comprised 49 (14%) females and 294 (86%) males, while the control group included 131 (38%) females and 213 (62%) males. As expected,^31^ sex distribution significantly differed between iRBD patients and controls (P < 0.0001). This difference did not impact analyses, as all comparisons were either stratified by sex or performed on W-scored measurements, which use controls as the reference group. iRBD patients did not differ significantly in age from controls (P = 0.43) but had higher MDS-UPDRS-III scores (P < 0.001) and lower MoCA scores (P < 0.001) (Table 1). Within the iRBD group, no significant differences between females and males were observed for age (P = 0.42), MDS-UPDRS-III scores (5.5 ±5.2 in females vs. 6.7 ±6.0 in males, P = 0.22) or MoCA scores (25.6 ± 3.9 in females vs. 25.5 ± 3.0 in males, P = 0.83) (Table 1).

**Table 1.** Demographics and clinical characteristics of participants.

| Variables | iRBD (n=343) |  |  | Controls (n=344) |  |  | P-value |  |  |
| --- | --- | --- | --- | --- | --- | --- | --- | --- | --- |
|  | Total | Females | Males | Total | Females | Males | Total<sup>a</sup> | iRBD<sup>b</sup> | Controls<sup>c</sup> |
| Sex, n (%) | 343 | 49 (14%) | 294 (86%) | 344 | 131 (38%) | 213 (62%) | <0.0001<sup>d</sup> | - | - |
| Age, years | 67.0±6.9 | 66.3±5.9 | 67.2±7.1 | 66.6±6.9 | 66.3±5.9 | 67.0±6.9 | 0.43<sup>e</sup> | 0.42<sup>e</sup> | 0.16<sup>e</sup> |
| MDS-UPDRS-III | 6.5±5.9 | 5.5±5.2 | 6.7±6.0 | 3.1±5.8 | 4.0±8.3 | 2.6±3.7 | <0.001<sup>e</sup> | 0.22<sup>e</sup> | 0.16<sup>e</sup> |
| MoCA | 25.5±3.1 | 25.6±3.9 | 25.5±3.0 | 27.0±2.3 | 27.4±2.5 | 26.8±2.2 | <0.001<sup>e</sup> | 0.83<sup>e</sup> | 0.07<sup>e</sup> |

Data are presented as mean ± SD.
<sup>a</sup> iRBD patients versus controls.
<sup>b</sup> Female vs. male iRBD patients.
<sup>c</sup> Female vs. male controls.
<sup>d</sup> Chi-squared test.
<sup>e</sup> Independent two-sample t-test.
iRBD = isolated REM sleep behavior disorder; MDS = Movement Disorder Society; MoCA = Montreal Cognitive Assessment; SD = standard deviation; UPDRS-III = Unified Parkinson's Disease Rating Scale part III.

### Sex impacts cortical thickness differently in iRBD and controls

Before examining sex effects on cortical atrophy (i.e., reductions in cortical thickness beyond what is expected for age) in iRBD, we first assessed whether sex differences in cortical thickness varied between iRBD patients and controls. Vertex-wise surface analysis of cortical thickness revealed a significant sex-by-group interaction in three cortical clusters (Figure 1A, Table 2). The peaks were located in the left posterior cingulate cortex (7846 mm^2^, 16820 vertices, Talairach coordinates: x = –4.7, y = –27.9, z = 34.6, P = 0.0007) and superior parietal cortex (3279 mm^2^, 8099 vertices, x = –32.0, y = –38.9, z = 39.2, P = 0.0004), extending to sensorimotor and medial frontal cortex, and in the right paracentral cortex (8607 mm^2^, 18097 vertices, x = 5.2, y = –25.1, z = 66.3, P = 0.0004), extending to the motor cortex, medial frontal cortex, and dorsolateral prefrontal cortex (Figure 1A, Table 2). In the left posterior cingulate and right paracentral clusters, males with iRBD showed significantly reduced cortical thickness compared to females, while no significant sex differences were observed in controls (Figure 1B, Table S1). In contrast, in the left superior parietal cluster, males exhibited reduced cortical thickness compared to females in both groups, with this reduction being significantly more pronounced in iRBD patients (Figure 1B). Vertex-based analyses conducted separately within each group revealed similar clusters in iRBD, identifying 6 clusters (2 in the left hemisphere, 4 in the right hemisphere) where males had significantly lower cortical thickness than females (Figure S1, Table S2). In contrast, no significant male-female difference was detected in the control group. To verify the sex effect on other structural measures, vertex-based cortical surface area and volume analyses were conducted. Results revealed a significant sex-group interaction on cortical surface area and cortical volume in the right superior frontal cortex (Figure S2A, Table S3). When conducting analyses separately in iRBD and controls, iRBD males showed reduced cortical surface area and volume in the right superior frontal cortex compared to females (Figure S2B, Table S3). This effect was not present in controls. Taken together, these findings support a significant interaction between sex and group, indicating that males with iRBD present greater cortical thinning than females, with similar age and clinical severity.

**Figure 1.**
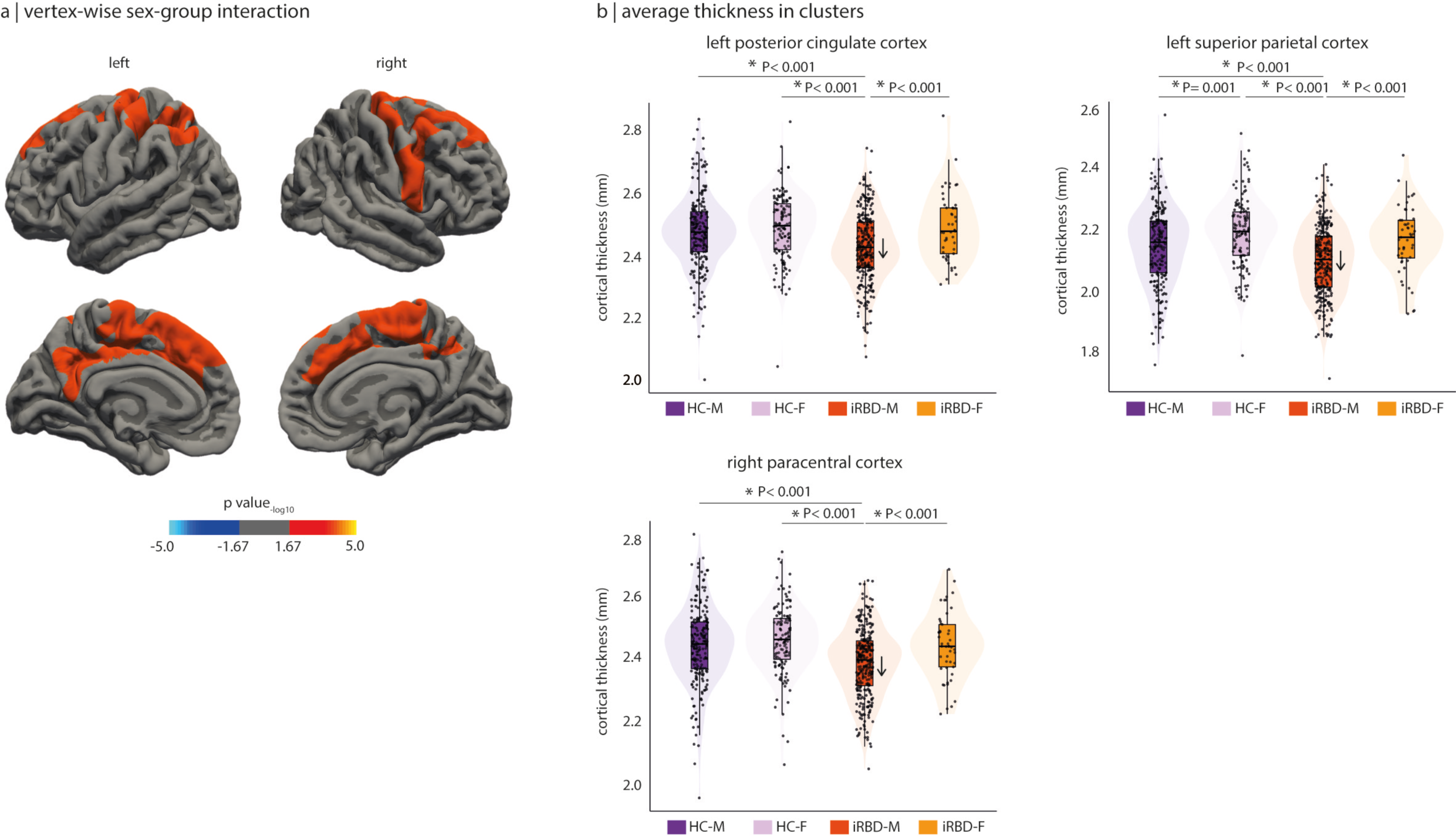
Vertex-wise cortical thickness analysis reveals significant sex-group interaction. (a) Clusters showing significant sex-group interaction on cortical thickness. The colour bar indicates the statistical significance on a logarithmic scale of *P*-values (−log10), with positive values showing significant decreases in iRBD males compared to iRBD females and controls. (b) Average cortical thickness (in mm) across groups in significant clusters, showing significant reduction in cortical thickness in iRBD males compared to iRBD females and controls. The P-values indicate significant differences between groups after conducting independent two sample t-tests. See Table S1 for details. HC-M = healthy controls males; HC-F = healthy controls females; iRBD-M = iRBD males; iRBD-F= iRBDfemales.

**Table 2.** Vertex-based cortical thickness analyses of sex effects.

| Cluster peak location<sup>a</sup> | Cluster size, mm<sup>2</sup> | Number of vertices | Talairach coordinates |  |  | −log<sub>10</sub> *P*-value |
| --- | --- | --- | --- | --- | --- | --- |
|  |  |  | *x* | *y* | *z* |  |
| Sex-by-group interaction |  |  |  |  |  |  |
| Left posterior cingulate cortex | 7846 | 16820 | −4.7 | −27.9 | 34.6 | 3.18 |
| Left superior parietal cortex | 3279 | 8099 | −32.0 | −38.9 | 39.2 | 3.44 |
| Right paracentral cortex | 8607 | 18097 | 5.2 | −25.1 | 66.3 | 3.40 |
| Sex effect in iRBD |  |  |  |  |  |  |
| Left postcentral cortex | 13676 | 29 086 | −34.5 | −32.9 | 41.1 | 5.53 |
| Left inferior parietal cortex | 2616 | 4909 | −44.6 | −73.2 | 14.4 | 4.26 |
| Right caudal middle frontal cortex | 3415 | 6690 | 37.6 | 2.3 | 34.7 | 3.89 |
| Right inferior parietal cortex | 3276 | 7191 | 41.6 | −70.3 | 36.0 | 3.16 |
| Right rostral middle frontal cortex | 2778 | 4435 | 22.2 | 52.1 | 24.1 | 2.81 |
| Right paracentral cortex | 2448 | 5660 | 7.8 | −11.9 | 60.6 | 4.09 |

Clusters were significant after Monte-Carlo simulation, with cluster- and vertex-level P-values set at $P < 0.05$ .
<sup>a</sup> Only the region of the peak vertex is indicated (see Figure 1 for mapping).
iRBD = isolated REM sleep behavior disorder.

### Cortical atrophy differs based on sex in iRBD

We next examined whether the pattern and extent of cortical atrophy differed between females and males with iRBD using a parcel-wise approach. To address this, we parcellated the cortical surface into 1000 cortical regions across hemispheres, extracted cortical thickness values, harmonized them across acquisition sites, and standardized them for age and sex using regression models from healthy controls (W-scores). Cortical atrophy in iRBD patients was assessed using one-sample t-tests, determining whether regional values significantly deviated from 0, corresponding to expected value in age– and sex-matched controls (i.e., no atrophy compared to controls). Significant negative deviations indicated cortical thinning in iRBD patients. In the left hemisphere, iRBD males exhibited significant atrophy (P < 0.05) in 191 regions (38%), predominantly affecting the sensorimotor cortex, perisylvian region, and occipital cortex (Figure 2A). Among these, 183 regions (37%) remained significant after FDR correction, with t-scores ranging from –2.3 to –12.1. In contrast, iRBD females exhibited atrophy in 53 regions (11%) regions, largely overlapping with affected regions in males, particularly in the sensorimotor and occipital cortices. However, only 4 regions (1%) remained significant after FDR correction, with t-scores ranging from –3.7 to –4.8 (Figure 2A). In the right hemisphere, iRBD males exhibited atrophy in 298 regions (60%) (P < 0.05), primarily affecting the sensorimotor cortex, perisylvian region, and occipitoparietal cortex. After FDR correction, 262 regions (53%) remained significant, with t-scores ranging between –2.2 to –6.7 (Figure 2B). In iRBD females, 84 regions (17%) showed significant thinning (P < 0.05), again in similar areas affected in males; however, none remained significant after FDR correction. These findings reveal that cortical atrophy in iRBD in markedly less widespread and less severe in females compared to males (χ^2^ = 210.65, P < 0.0001), despite comparable age and clinical severity. However, the spatial pattern of atrophy is largely similar between sex, suggesting the presence of sex-specific protective mechanisms that may mitigate brain neurodegeneration in females.

**Figure 2.**
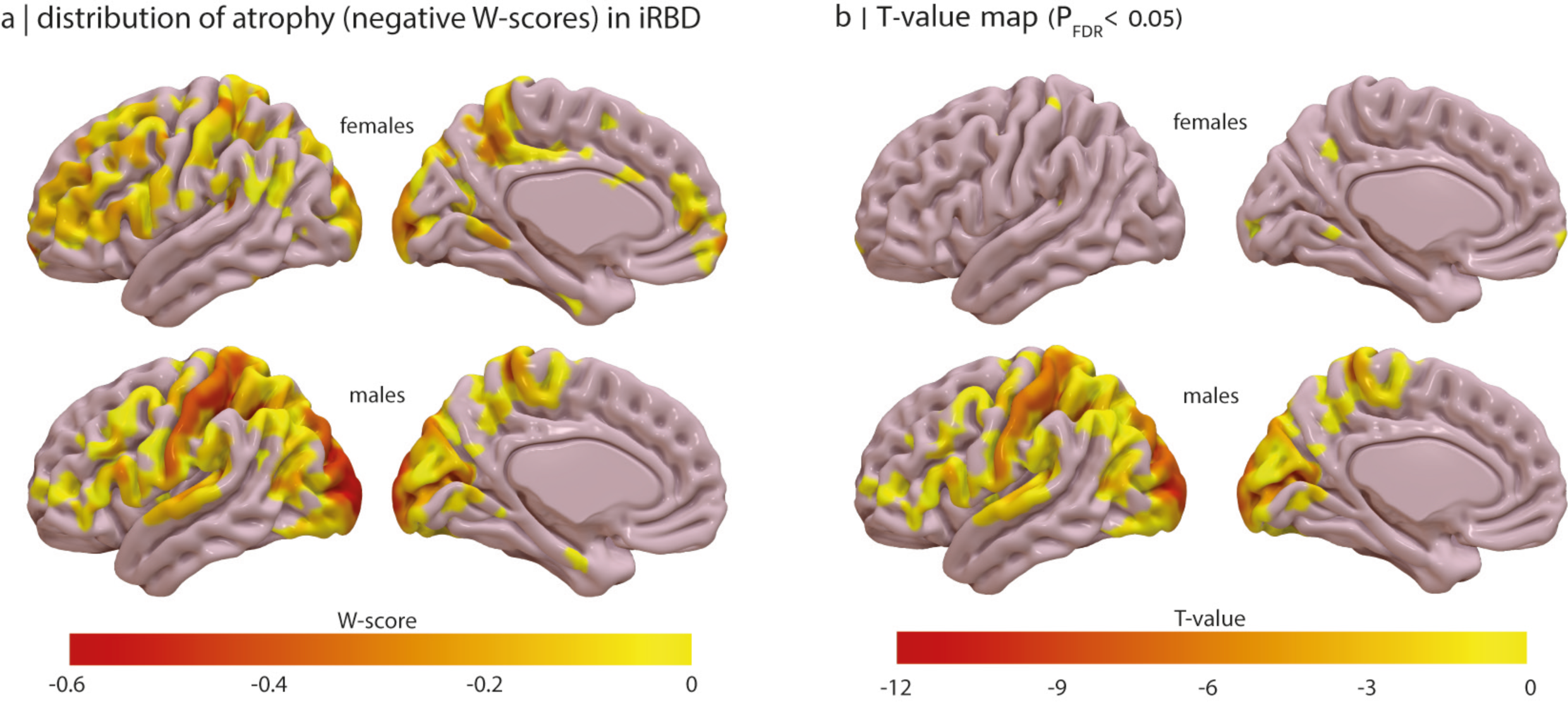
Brain map showing atrophy patterns stratified by sex in iRBD. (a) distribution of reduced cortical thickness in females and males with iRBD. The colour bar indicates the range of negative W-score describing cortical atrophy compared to controls. (b) FDR corrected significantly atrophied regions in females and males with iRBD respectively. The colour bar indicates the range of t-values for each regions showing significant atrophy in iRBD. FDR= false discovery rate.

### Oestrogen-related receptor genes underly sex effects in iRBD

To investigate the mechanisms potentially protecting iRBD females from cortical atrophy, we examined whether brain regions with less thinning in females expressed distinct gene expression patterns that may confer neuroprotection. We first tested whether the spatial distribution of gene expression was associated with sex differences in cortical atrophy. Using W-scored cortical thickness values, we applied a linear regression model to calculate a sex interaction estimate for each parcellated region in the left hemisphere, as gene expression data was primarily available for this hemisphere. A positive sex interaction coefficient indicated regions where iRBD females exhibited less cortical thinning than males.

PLS regression identified two significant latent variables (LV1 and LV3) that explained significantly more covariance between gene expression and sex interaction estimates than spatially constrained null models. LV1 explained 19% of the covariance (compared to 11% in null models, P = 0.007) and LV3 explained 11% (compared to 6% in null models, P = 0.02) (Figure 3A). The regional weights attributed to these significant latent variables were positively correlated with sex interaction estimates on cortical thickness (LV1: r = 0.44, P < 0.0001; LV3: r = 0.33, P < 0.0001) (Figure 3B-3C), indicating that genes positively weighted on these latent variables were overexpressed in regions where iRBD females had less cortical thinning than males.

**Figure 3.**
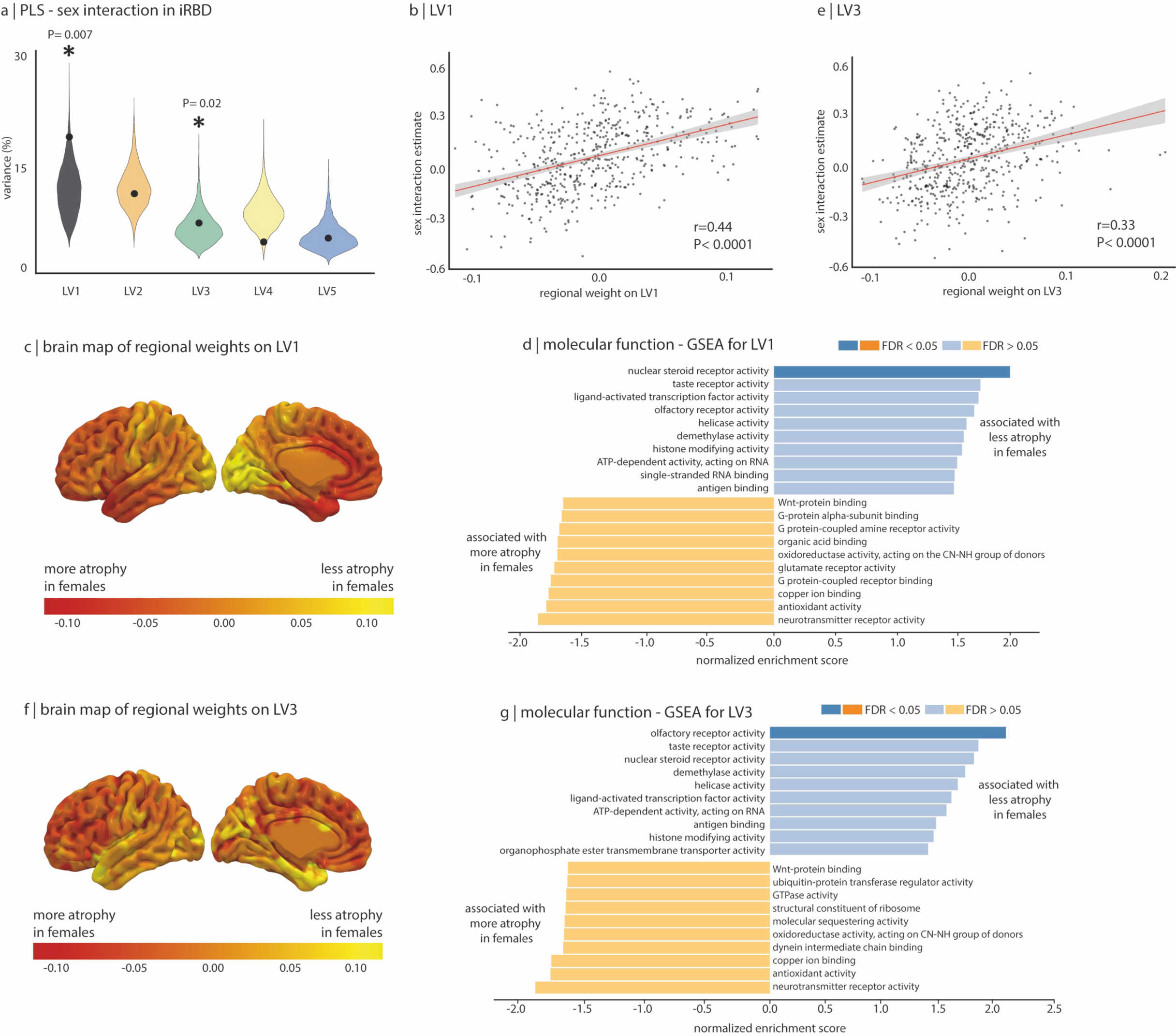
GSEA on sex interaction estimates of cortical thickness in iRBD. (a) Violin plots showing the percentage of variance in sex interaction estimates of cortical thickness explained by gene expression; the dot represents the empirical variance, and the asterisk indicates the components that were significant against spatial null models. (b) Scatterplot of the association between the sex interaction estimates and the regional weights of the first and third (c) latent variables. (d) Brain maps of the sex interaction coefficients and their regional weights of the first and third (f) latent variables. (e)-(g) The top 10 molecular function terms from the Gene Ontology Consortium knowledge base that are enriched in the positively and negatively weighted gene sets associated with sex effect on cortical thickness in iRBD. Terms are ranked based on the normalized enrichment score; darker coloured bars present significantly enriched terms after FDR correction. PLS= Partial least square; LV= latent variable; FDR = false discovery rate.

To identify molecular functions associated with cortical atrophy resilience in iRBD females, we ranked gene based on bootstrapping, estimating the robustness of each gene’s contribution to LV1 and LV3. For LV1, gene set enrichment analysis revealed significant enrichment in nuclear steroid receptor activity (8/20 genes, 40%; normalized enrichment score-NES=2.00, PFDR = 0.027), which was associated with less cortical thinning in iRBD females (Figure 3D-3E, Table 3). The most strongly associated genes in this function were *ESRRG* (bootstrap ratio = 13.3) and *ESRRA* (bootstrap ratio = 10.9), encoding for estrogen-related receptor gamma and alpha, followed various nuclear receptor subfamily group members (*NR3C2, NR2C1, NR3C1*) and by *PPARD* (bootstrap ratio = 7.26), encoding for peroxisome proliferator-activated receptor delta (Table 4). For the LV3, gene set enrichment analysis also uncovered a molecular function significantly associated with positive sex interaction estimates, corresponding to reduced cortical thinning in iRBD females (Figure 3F). The most strongly associated gene terms were enriched in olfactory receptor activity (29/48 genes, 60%; NES= 2.08, PFDR = 0.015) (Figure 3G, Tables 5 and S4). No significant biological process or cellular component terms were associated with LV1 or LV3.

**Table 3.**
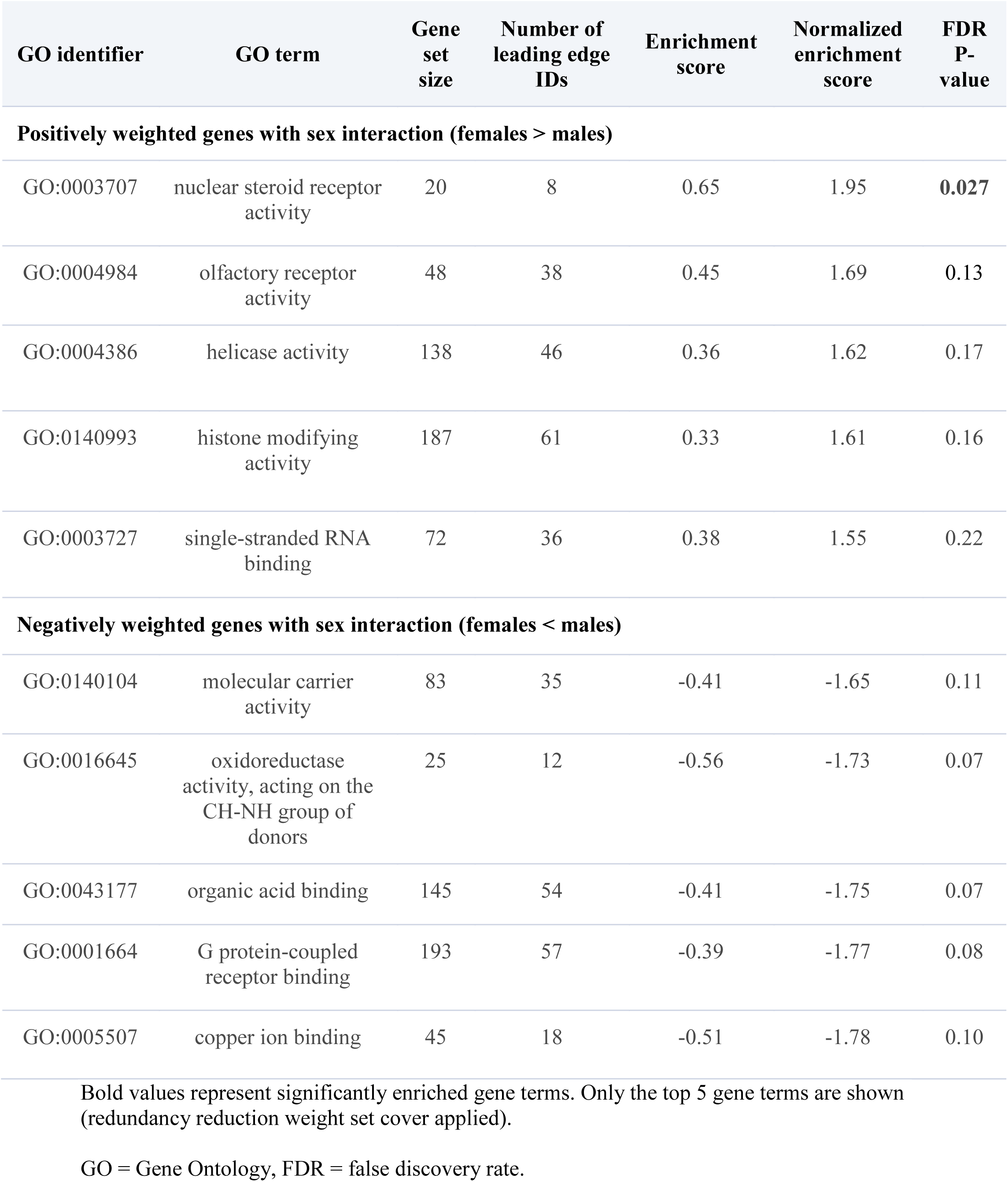
Molecular functions enriched in regions with sex interaction for LV1.

**Table 4.** Gene association within nuclear steroid receptor activity for LV1.

| Gene | Bootstrap ratio | Gene symbol | Gene name |
| --- | --- | --- | --- |
| ESRRG | 13.33 | *ESRRG* | Oestrogen-related receptor gamma |
| ESRRA | 10.92 | *ESRRA* | Oestrogen-related receptor alpha |
| NR3C2 | 9.07 | *NR3C2* | Nuclear receptor subfamily 3 group C member 2 |
| NR2C1 | 7.57 | *NR2C1* | Nuclear receptor subfamily 2 group C member 1 |
| NR3C1 | 7.31 | *NR3C1* | Nuclear receptor subfamily 3 group C member 1 |
| PPARD | 7.26 | *PPARD* | Peroxisome proliferator activated receptor delta |
| NR1D1 | 6.33 | *NR1D1* | Nuclear receptor subfamily 1 group D member 1 |
| ESRRB | 5.15 | *ESRRB* | Oestrogen-related receptor beta |

Only showing the top 8 out of 20 gene terms included in this molecular function.

**Table 5.** Molecular functions enriched in regions with sex interaction for LV3.

| GO identifier | GO term | Gene set size | Number of leading edge IDs | Enrichment score | Normalized enrichment score | FDR P-value |
| --- | --- | --- | --- | --- | --- | --- |
| **Positively weighted genes with sex interaction (females > males)** |  |  |  |  |  |  |
| GO:0004984 | olfactory receptor activity | 48 | 29 | 0.49 | 2.06 | **0.015** |
| GO:0003707 | nuclear steroid receptor activity | 20 | 8 | 0.54 | 1.80 | **0.09** |
| GO:0004386 | helicase activity | 138 | 53 | 0.33 | 1.67 | 0.11 |
| GO:0140993 | histone modifying activity | 187 | 53 | 0.27 | 1.46 | 0.27 |
| **Negatively weighted genes with sex interaction (females < males)** |  |  |  |  |  |  |
| GO:0003735 | structural constituent of ribosome | 156 | 65 | -0.37 | -1.60 | 0.10 |
| GO:0001664 | G protein-coupled receptor binding | 193 | 80 | -0.37 | -1.62 | 0.10 |
| GO:0016209 | antioxidant activity | 63 | 23 | -0.46 | -1.71 | 0.09 |
| GO:0043177 | organic acid binding | 145 | 51 | -0.41 | -1.75 | 0.09 |
| GO:0031681 | G-protein beta-subunit binding | 19 | 11 | -0.59 | -1.76 | 0.10 |
| GO:0030594 | neurotransmitter receptor activity | 71 | 26 | -0.47 | -1.82 | 0.08 |

Bold values represent significantly enriched gene terms. Only the top 5 gene terms are shown (redundancy reduction weight set cover applied).
GO = Gene Ontology, FDR = false discovery rate.

To verify that these findings were not driven by gene ontology platform selection, we conducted a secondary enrichment analysis using GOrilla.^62^ For LV1, steroid hormone receptor activity (PFDR = 0.005, enrichment score = 14.3) and nuclear steroid receptor activity (PFDR = 0.01, enrichment score = 6.1) emerged as the top enriched functions (Table S5), both including *ESRRG* and *ESRRA* (Tables S6 and S7). Interestingly, another common gene that was listed in both analyses from the different platforms was the peroxisome proliferator-activated receptor delta genes (*PPARD)*. No significant biological process or cellular component terms were identified for LV1 or LV3. Taken together, these results suggest that regions where iRBD females show less cortical thinning overexpress genes enriched in nuclear steroid receptors, particularly estrogen-related receptors (*ESRRG* and *ESRRA*).

### Brain-enriched expression of ESRRG in sex-related cortical atrophy

To further explore the systemic roles and biological relevance of *ESRRG* and *ESRRA* in sex-related cortical atrophy in iRBD, we analyzed their tissue-specific expression patterns using GTEx data. The GTEx dataset provides gene expression profiles from 54 non-diseased tissue types, collected from nearly 1,000 post-mortem donors^63^ enabling a comprehensive investigation of how these genes are expressed across human tissues.

The results revealed distinct expression profiles for *ESRRG* and *ESRRA* (Figure 4). Violin plots showed qualitative differences: *ESRRG* exhibited a more tissue-specific and brain-enriched expression pattern, whereas *ESRRA* showed a more ubiquitous distribution, with lower expression in the brain compared to other organs. Quantitative analysis confirmed these observations. Among tissues where each gene was overexpressed relative to the median expression across all tissues, *ESRRG* was significantly more enriched in brain tissues compared to *ESRRA* (13/27 (48%) tissue types for *ESRRG* vs. 2/27 (7%) for *ESRRA*, P = 0.002). Additionally, we analyzed the tissue expression patterns of *PPARD* following the same expression profiles of the 54 tissue types available from GTEx data. Results overall showed a ubiquitous distribution across tissue types, not brain specific compared to *ESRRG*. Details can be found in supplementary materials (Figure S3). Taken together, these findings highlight the more neural-specific relevance of *ESRRG*, suggesting its potential role in the selective resilience of brain regions to sex-related cortical atrophy in iRBD.

**Figure 4.**
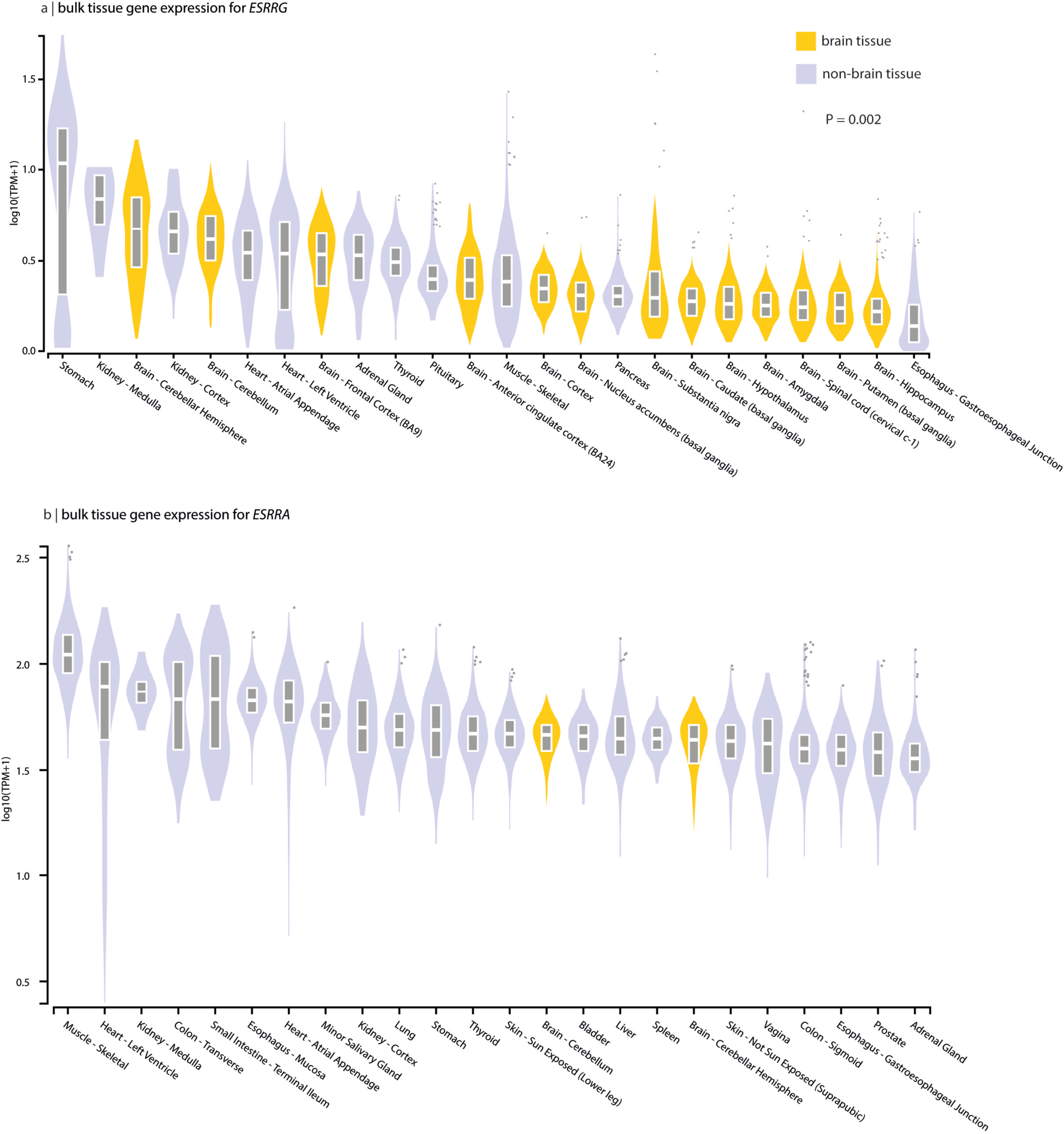
Top 24 tissues expression of *ESRRG* and *ESRRA*. (a) Violin plots of expression values of *ESRRG* across tissue types, showing a brain specific (yellow) expression pattern compared to other tissue types (lilac) (P<0.002). (b) Tissue expression values of *ESRRA*, showing a more ubiquitous distribution across tissue types (lilac), and not brain specific (yellow) pattern of expression. Box plots are shown as median, 25^th^ and 75^th^ percentiles. Points appearing outside of the plots corresponds to outliers above or below 1.5 times the interquartile range. TPM = transcripts per million.

## Discussion

This study investigated sex-related differences in cortical atrophy in iRBD, a prodromal stage of synucleinopathies, using a large multicenter MRI dataset of polysomnography-confirmed iRBD patients and controls. Our findings demonstrate that females with iRBD exhibit significantly less cortical thinning than males, despite similar age and clinical severity, with this effect absent in healthy controls. While both sexes share similar spatial patterns of cortical thinning, females exhibited more restricted and less severe atrophy. Transcriptomic analyses revealed that regions showing less cortical thinning in females, notably in occipital and sensorimotor areas, were enriched for nuclear steroid receptor functions, with estrogen-related genes *ESRRG* and *ESRRA* emerging as strongly associated, particularly *ESRRG*, which displayed a more brain-specific expression pattern. These findings suggest that selective resilience mechanisms, potentially mediated by estrogen-related pathways, contribute to reduced neurodegeneration in iRBD females compared to males.

Sex differences in manifest synucleinopathies have been widely reported,^15,16^ yet their presence in prodromal stages such as iRBD has not been well characterized. Here, we demonstrate that males with iRBD exhibit significantly greater cortical thinning than females, despite comparable age and clinical severity. Importantly, this sex difference was absent in control participants. Our findings align with studies in manifest DLB and PD, which consistently report greater neurodegeneration in males.^16,20,21,65,66^ In PD, vertex-based analyses have shown sex effects on cortical thickness and volume, particularly in regions overlapping those identified in our iRBD cohort, with males displaying greater cortical thinning and smaller volumes than females.^65^ Males with PD also exhibit more severe motor impairment and higher prevalence of RBD symptoms compared to females.^65^ Deformation-based morphometry and connectivity studies further support this trend, showing that females with PD exhibit less cortical atrophy and less white matter connectivity disruption than males.^67^ Similarly, Yadav et al. reported significant reduced cortical thickness and lower structural network correlations in males with PD compared to female patients.^20^ In early PD stages, sex-specific cortical connectivity patterns showed that females had increased connectivity, correlated with more preserved motor function over time.^66^ Similar findings have been reported in DLB, where volumetric analyses demonstrate that females exhibit less cortical grey matter loss than males,^22^ with differences particularly pronounced at younger ages.^22^ Given that iRBD represents a prodromal stage for PD and DLB, our findings suggest that similar sex-based neuroanatomical resilience mechanisms may already be present in iRBD females, contributing to less severe and more restricted cortical thinning compared to males.

Selective resilience refers to the ability of specific brain regions to resist neurodegeneration, even in the presence of pathological processes.^68^ In this study, only 1% of cortical regions in iRBD females showed significant atrophy, compared to 37% in males, despite both groups having similar age, clinical severity, and spatial patterns of cortical thinning, in the sensorimotor cortex, occipital cortex, and perisylvian areas. Cortical thinning was particularly prominent in the occipital cortex, but also in parietal and dorsolateral prefrontal regions, which is consistent with previous reports in iRBD,^7,8^ PD,^69^ and DLB,^22,70^ suggesting overlapping neurodegenerative mechanisms. To investigate the biological basis of selective resilience in iRBD females, we analyzed gene expression patterns in regions showing sex-based differences in cortical thinning. Gene enrichment analysis identified nuclear steroid receptor functions, particularly estrogen-related receptors (*ESRRG* and *ESRRA*) genes, as strongly associated with regions showing less atrophy in iRBD females. Notably, *ESRRG* exhibited more brain-specific overexpression. This contrasts with the more ubiquitous expression of *ESRRA*, underscoring its potential role in supporting brain resilience.

Estrogen-related receptors (ERRγ, ERRα, ERRβ) are nuclear receptors involved in mitochondrial biogenesis, energy homeostasis, and neuronal metabolism.^71^ ERRγ and ERRα act as co-regulators of PGC-1α, a key regulator of mitochondrial function, oxidative metabolism, and synaptic maintenance.^71,72^ PGC-1a is highly expressed in neurodegeneration-vulnerable regions, including the cerebral cortex, hippocampus, striatum, and substantia nigra.^73^ Disruptions in PGC-1α signaling and mitochondrial dysfunction are well-established contributors of synucleinopathy-related neurodegeneration, leading to energy deficits, oxidative stress, and impaired protein clearance.^74,75^ Work from our group has shown that atrophy-prone regions in iRBD overexpress mitochondrial-related genes,^12^ supporting the idea that neuronal energy deficits play a role in disease progression. Furthermore, recent findings indicate that *ESRRG* expression enhances mitochondrial function, protecting dopaminergic neurons from synuclein-induced toxicity and promoting resilience against neurodegeneration.^76^ Fox et al. demonstrated that deleting ERRγ in dopaminergic neurons increased vulnerability to alpha-synuclein toxicity, reduced mitochondrial gene expression, and decreased mitochondrial number, whereas ERRγ overexpression reduced alpha-synuclein aggregation and delayed neurodegeneration.^76^ Further supporting this hypothesis, Ciron et al. demonstrated that PGC-1α deficiency exacerbates neurodegeneration in PD, leading to increased mitochondrial dysfunction and alpha-synuclein toxicity, with males being more vulnerable to neurodegeneration following PGC-1α loss.^77^ Given that PGC-1α and ERRγ interact to regulate mitochondrial function, our findings suggest that *ESRRG* overexpression in atrophy-resistant cortical areas in iRBD females may contribute to mitochondrial protection, counteracting the neurodegenerative processes observed in males.

The neuroprotective potential of *ESRRG* is particularly compelling given our GTEx analysis, which revealed significant overexpression of *ESRRG* in the brain, in contrast to *ESRRA*, which is more ubiquitously expressed. McMeekin et al. showed that disabling *ESRRA* does not significantly impact mitochondrial gene expression, reinforcing the idea that neuronal mitochondrial regulation may be more strongly influenced by *ESRRG*.^78^ These findings suggest that *ESRRG* may play a brain-specific role in neuroprotection, contributing to greater resilience against neurodegeneration in iRBD females. Beyond *ESRRG* and *ESRRA*, another notable gene identified in atrophy-resistant regions of iRBD females was the peroxisome proliferator-activated receptor delta gene (*PPARD*). PPARδ is a nuclear receptor involved in lipid metabolism, energy homeostasis, and mitochondrial function.^79^ Among PPAR subtypes, PPARδ is the most highly expressed in the brain, where it regulates neuronal cell survival, neuroinflammation, and resistance to neurodegeneration resistance.^79,80^ PPARδ has demonstrated neuroprotective effects in multiple neurodegenerative diseases, including Parkinson’s disease,^81^ Huntington disease,^82^ and Alzheimer’s disease^83^. Preclinical and clinical studies suggest that PPARδ activation protects against mitochondrial dysfunction in PD models, reduces amyloid burden, and improves cognitive function in Alzheimer’s disease.^81,83,82^ The overrepresentation of *PPARD* in atrophy-resistant cortical regions of iRBD females, alongside *ESRRG*, suggests a broader network of transcription factors contributing to sex-based resilience and highlights potential therapeutic targets for synucleinopathies.

This study has some limitations. First, while the multicentric dataset is a major strength, providing a large and diverse sample, the availability of detailed clinical and demographic data, including sex versus gender distinctions, was limited. Future studies should aim to disentangle these variables to better explore brain-clinical relationships in iRBD. Second, the male predominance in iRBD resulted in a smaller proportion of female participants. Despite this limitation, our study provides robust evidence of sex-related differences, making it the largest neuroimaging investigation of females with iRBD to date. Third, the gene expression data used in this study, obtained from the Allen Human Brain Atlas, provided high-resolution insights into over 20,000 genes across brain regions. However, these data were derived from healthy postmortem brains rather than iRBD-specific samples. Nevertheless, the ratios of males and females in the donors (83%) was equivalent to the one in our iRBD group (83%). These samples represent a good estimate of the gene expression in different brain regions before significant neurodegeneration occurs. Collecting postmortem gene expression data for every brain region remains a colossal challenge, particularly for iRBD, where most patients convert to a neurodegenerative disease by the time their brains become available for study. Lastly, this study provides cross-sectional insights into sex differences in iRBD. Longitudinal follow-ups will be essential to understanding how sex-based differences influence phenoconversion to PD, DLB or MSA. As more patients progress to manifest synucleinopathies, future analyses will enable sex-stratified investigations into disease progression.

In summary, this study reveals significant sex-related differences in cortical atrophy in iRBD, with females showing less severe and less widespread cortical thinning than males. Gene enrichment analyses identified estrogen-related pathways, particularly the *ESRRG* gene, as potential contributors to selective resilience in females. These findings provide new insights into sex-based neuroprotection in prodromal synucleinopathies, highlighting promising directions for targeted therapeutic strategies. the

## Data Availability

The data used in this study were obtained from multiple collaborating centres, each of which retains ownership of their respective datasets. The principal investigator had authorized access to all data necessary for the analyses performed in this study. However, the accessibility and sharing of data are subject to the local policies and restriction criteria of each centre involved. As such, data availability is restricted, and requests for access should be directed to the respective institutions, pending their specific data access and sharing guidelines.

## ACKNOWLEDGEMENT

The authors thank all participants included in the different cohorts studied. M.F. holds the Merit scholarship from the Faculty of Medicine of University of Montreal and the Bourse d’excellence Solange-Saul. S.R. reports a Junior 1 research scholar award from Fonds de recherche du Québec – Santé.

## Appendix 1

List of the contributors involved in the ICEBERG Study Group:

Steering committee: Marie Vidailhet, MD, PhD, (Pitié-Salpêtrière Hospital, Paris, principal investigator of ICEBERG), Jean-Christophe Corvol, MD, PhD (Pitié-Salpêtrière Hospital, Paris, scientific lead), Isabelle Arnulf, MD, PhD (Pitié-Salpêtrière Hospital, Paris, member of the steering committee), Stéphane Lehericy, MD, PhD (Pitié-Salpêtrière Hospital, Paris, member of the steering committee);

Clinical data: Marie Vidailhet, MD, PhD, (Pitié-Salpêtrière Hospital, Paris, coordination), Graziella Mangone, MD, PhD (Pitié-Salpêtrière Hospital, Paris, co-coordination), Jean-Christophe Corvol, MD, PhD (Pitié-Salpêtrière Hospital, Paris), Isabelle Arnulf, MD, PhD (Pitié-Salpêtrière Hospital, Paris), Smaranda Leu MD (Pitié-Salpêtrière Hospital, Paris), Sara Sambin, MD (Pitié-Salpêtrière Hospital, Paris), Jonas Ihle, MD (Pitié-Salpêtrière Hospital, Paris), Caroline Weill, MD, (Pitié-Salpêtrière Hospital, Paris), Poornima MENON MD, (Pitié-Salpêtrière Hospital, Paris), David Grabli, MD, PhD (Pitié-Salpêtrière Hospital, Paris); Florence Cormier-Dequaire, MD (Pitié-Salpêtrière Hospital, Paris); Louise Laure Mariani, MD, PhD (Pitié-Salpêtrière Hospital, Paris), Emmanuel Roze, MD, PhD, (Pitié-Salpêtrière Hospital, Paris), Cécile Delorme, MD (Pitié-Salpêtrière Hospital, Paris), Elodie Hainque MD, PhD, (Pitié-Salpêtrière Hospital, Paris), Aurelie Méneret (MD, PhD, (Pitié-Salpêtrière Hospital, Paris), Bertrand Degos, MD, PhD (Avicenne Hospital, Bobigny); Neuropsychological data: Richard Levy, MD (Pitié-Salpêtrière Hospital, Paris, coordination), Fanny Pineau, MS (Pitié-Salpêtrière Hospital, Paris, neuropsychologist), Julie Socha, MS (Pitié-Salpêtrière Hospital, Paris, neuropsychologist), Eve Benchetrit, MS (La Timone Hospital, Marseille, neuropsychologist), Virginie Czernecki, MS (Pitié-Salpêtrière Hospital, Paris, neuropsychologist), Marie-Alexandrine Glachant, MS (Pitié-Salpêtrière Hospital, Paris, neuropsychologist);

Eye movement: Sophie Rivaud-Pechoux, PhD (ICM, Paris, coordination); Elodie Hainque, MD, PhD (Pitié-Salpêtrière Hospital, Paris);

Sleep assessment: Isabelle Arnulf, MD, PhD (Pitié-Salpêtrière Hospital, Paris, coordination), Smaranda Leu Semenescu, MD (Pitié-Salpêtrière Hospital, Paris), Pauline Dodet, MD (Pitié-Salpêtrière Hospital, Paris);

Genetic data: Jean-Christophe Corvol, MD, PhD (Pitié-Salpêtrière Hospital, Paris, coordination), Graziella Mangone, MD, PhD (Pitié-Salpêtrière Hospital, Paris, co-coordination), Samir Bekadar, MS (Pitié-Salpêtrière Hospital, Paris, biostatistician), Alexis Brice, MD (ICM, Pitié-Salpêtrière Hospital, Paris), Suzanne Lesage, PhD (INSERM, ICM, Paris, genetic analyses);

Metabolomics: Fanny Mochel, MD, PhD (Pitié-Salpêtrière Hospital, Paris, coordination), Farid Ichou, PhD (ICAN, Pitié-Salpêtrière Hospital, Paris), Vincent Perlbarg, PhD, Pierre and Marie Curie University), Benoit Colsch, PhD (CEA, Saclay), Arthur Tenenhaus, PhD (Supelec, Gif-sur-Yvette, data integration);

Brain MRI data: Stéphane Lehericy, MD, PhD (Pitié-Salpêtrière Hospital, Paris, coordination), Rahul Gaurav, MS, (Pitié-Salpêtrière Hospital, Paris, data analysis), Nadya Pyatigorskaya, MD, PhD, (Pitié-Salpêtrière Hospital, Paris, data analysis); Lydia Yahia-Cherif, PhD (ICM, Paris, Biostatistics), Romain Valabregue, PhD (ICM, Paris, data analysis), Cécile Galléa, PhD (ICM, Paris);

DaTscan imaging data: Marie-Odile Habert, MCU-PH (Pitié-Salpêtrière Hospital, Paris, coordination);

Voice recording: Dijana Petrovska, PhD (Telecom Sud Paris, Evry, coordination), Laetitia Jeancolas, MS (Telecom Sud Paris, Evry);

Study management: Vanessa Brochard (Pitié-Salpêtrière Hospital, Paris, coordination), Alizé Chalançon (Pitié-Salpêtrière Hospital, Paris, Project manager), Carole Dongmo-Kenfack (Pitié-Salpêtrière Hospital, Paris, clinical research assistant); Christelle Laganot (Pitié-Salpêtrière Hospital, Paris, clinical research assistant), Valentine Maheo (Pitié-Salpêtrière Hospital, Paris, clinical research assistant), Manon Gomes (Pitié-Salpêtrière Hospital, Paris, clinical research assistant).

## References

1. Hu, M. T. REM sleep behavior disorder (RBD). Neurobiol. Dis. 143, 104996 (2020).

2. American Academy of Sleep Medicine. The International Classification of Sleep Disorders — Third Edition (ICSD-3). (American Academy of Sleep Medicine, 2014).

3. Galbiati, A., Verga, L., Giora, E., Zucconi, M. & Ferini-Strambi, L. The risk of neurodegeneration in REM sleep behavior disorder: A systematic review and meta-analysis of longitudinal studies. Sleep Med. Rev. 43, 37–46 (2019).

4. Postuma, R. B. et al. Risk and predictors of dementia and parkinsonism in idiopathic REM sleep behaviour disorder: a multicentre study. Brain 142, 744–759 (2019).

5. Miglis, M. G. et al. Biomarkers of conversion to α-synucleinopathy in isolated rapid-eye-movement sleep behaviour disorder. Lancet Neurol. 20, 671–684 (2021).

6. Rahayel, S. et al. Patterns of cortical thinning in idiopathic rapid eye movement sleep behavior disorder. Mov. Disord. 30, 680–687 (2015).

7. Campabadal, A. et al. Cortical gray matter progression in idiopathic REM sleep behavior disorder and its relation to cognitive decline. NeuroImage Clin. 28, 102421 (2020).

8. Rahayel, S. et al. Brain atrophy in prodromal synucleinopathy is shaped by structural connectivity and gene expression. Brain 145, 3162–3178 (2022).

9. Rahayel, S. et al. Abnormal Gray Matter Shape, Thickness, and Volume in the Motor Cortico-Subcortical Loop in Idiopathic Rapid Eye Movement Sleep Behavior Disorder: Association with Clinical and Motor Features. Cereb. Cortex 28, 658–671 (2018).

10. Rahayel, S. et al. Cortical and subcortical gray matter bases of cognitive deficits in REM sleep behavior disorder. Neurology 90, (2018).

11. Rahayel, S. et al. A Prodromal Brain-Clinical Pattern of Cognition in Synucleinopathies. Ann. Neurol. 89, 341–357 (2021).

12. Rahayel, S. et al. Mitochondrial function-associated genes underlie cortical atrophy in prodromal synucleinopathies. Brain 146, 3301–3318 (2023).

13. Vegeto, E. et al. The Role of Sex and Sex Hormones in Neurodegenerative Diseases. Endocr. Rev. 41, 273–319 (2019).

14. Elliott, J. E. et al. Baseline characteristics of the North American prodromal Synucleinopathy cohort. Ann. Clin. Transl. Neurol. 10, 520–535 (2023).

15. Raheel, K. et al. Sex differences in alpha-synucleinopathies: a systematic review. Front. Neurol. 14, (2023).

16. Chiu, S. Y. et al. Sex differences in dementia with Lewy bodies: Focused review of available evidence and future directions. Parkinsonism Relat. Disord. 107, 105285 (2023).

17. Georgiev, D., Hamberg, K., Hariz, M., Forsgren, L. & Hariz, G.-M. Gender differences in Parkinson’s disease: A clinical perspective. Acta Neurol. Scand. 136, 570–584 (2017).

18. Choudhury, P. et al. The temporal onset of the core features in dementia with Lewy bodies. Alzheimers Dement. 18, 591–601 (2022).

19. Kang, K. W., Choi, S.-M. & Kim, B. C. Gender differences in motor and non-motor symptoms in early Parkinson disease. Medicine (Baltimore) 101, e28643 (2022).

20. Yadav, S. K. et al. Gender-based analysis of cortical thickness and structural connectivity in Parkinson’s disease. J. Neurol. 263, 2308–2318 (2016).

21. Tremblay, C. et al. Sex effects on brain structure in de novo Parkinson’s disease: a multimodal neuroimaging study. Brain 143, 3052–3066 (2020).

22. Oltra, J. et al. Sex differences in brain atrophy in dementia with Lewy bodies. Alzheimers Dement. 20, 1815–1826 (2024).

23. Picillo, M. et al. The relevance of gender in Parkinson’s disease: a review. J. Neurol. 264, 1583–1607 (2017).

24. Ventura-Clapier, R. et al. Mitochondria: a central target for sex differences in pathologies. Clin. Sci. 131, 803–822 (2017).

25. Ullah, M. F. et al. Impact of sex differences and gender specificity on behavioral characteristics and pathophysiology of neurodegenerative disorders. Neurosci. Biobehav. Rev. 102, 95–105 (2019).

26. Bourque, M., Morissette, M. & Di Paolo, T. Neuroactive steroids and Parkinson’s disease: Review of human and animal studies. Neurosci. Biobehav. Rev. 156, 105479 (2024).

27. Litim, N., Morissette, M. & Di Paolo, T. Neuroactive gonadal drugs for neuroprotection in male and female models of Parkinson’s disease. Neurosci. Biobehav. Rev. 67, 79–88 (2016).

28. Calabresi, P. et al. Alpha-synuclein in Parkinson’s disease and other synucleinopathies: from overt neurodegeneration back to early synaptic dysfunction. Cell Death Dis. 14, 1– 16 (2023).

29. Li, X. et al. Sex differences in rapid eye movement sleep behavior disorder: A systematic review and meta-analysis. Sleep Med. Rev. 71, 101810 (2023).

30. Castelnuovo, A., Marelli, S., Mombelli, S., Salsone, M. & Ferini-Strambi, L. Idiopathic RBD: the role of gender. J. Neurol. 267, 2157–2158 (2020).

31. Zhou, J. et al. Gender differences in REM sleep behavior disorder: a clinical and polysomnographic study in China. Sleep Med. 16, 414–418 (2015).

32. Mahale, R. R., Yadav, R. & Pal, P. Kr. Rapid eye movement sleep behaviour disorder in women with Parkinson’s disease is an underdiagnosed entity. J. Clin. Neurosci. 28, 43– 46 (2016).

33. Fernández-Arcos, A., Iranzo, A., Serradell, M., Gaig, C. & Santamaria, J. The Clinical Phenotype of Idiopathic Rapid Eye Movement Sleep Behavior Disorder at Presentation: A Study in 203 Consecutive Patients. Sleep 39, 121–132 (2016).

34. Mano, M., Nomura, A. & Sasanabe, R. Gender Difference in REM Sleep Behavior Disorder in Japanese Population: Polysomnography and Sleep Questionnaire Study. J. Clin. Med. 13, 914 (2024).

35. Zhang, H. et al. Risk Factors for Phenoconversion in RAPID EYE MOVEMENT Sleep Behavior Disorder. Ann. Neurol. 91, 404–416 (2022).

36. Rahayel, S. et al. ^99m^ Tc-HMPAO SPECT Perfusion Signatures Associated With Clinical Progression in Patients With Isolated REM Sleep Behavior Disorder. Neurology 102, e208015 (2024).

37. Marek, K. et al. The Parkinson’s progression markers initiative (PPMI) – establishing a PD biomarker cohort. Ann. Clin. Transl. Neurol. 5, 1460–1477 (2018).

38. Joza, S. et al. Distinct brain atrophy progression subtypes underlie phenoconversion in isolated REM sleep behaviour disorder. Preprint at 10.1101/2024.09.05.24313131 (2024).

39. McKeith, I. G. et al. Diagnosis and management of dementia with Lewy bodies: Fourth consensus report of the DLB Consortium. Neurology 89, 88–100 (2017).

40. Postuma, R. B. et al. MDS clinical diagnostic criteria for Parkinson’s disease: MDS-PD Clinical Diagnostic Criteria. Mov. Disord. 30, 1591–1601 (2015).

41. Wenning, G. K. et al. The Movement Disorder Society Criteria for the Diagnosis of Multiple System Atrophy. Mov. Disord. 37, 1131–1148 (2022).

42. Goetz, C. G. et al. Movement Disorder Society-sponsored revision of the Unified Parkinson’s Disease Rating Scale (MDS-UPDRS): Scale presentation and clinimetric testing results. Mov. Disord. 23, 2129–2170 (2008).

43. Nasreddine, Z. S., et al. The Montreal Cognitive Assessment, MoCA: A Brief Screening Tool For Mild Cognitive Impairment. J. Am. Geriatr. Soc. 53, 695–699 (2005).

44. Monereo-Sánchez, J. et al. Quality control strategies for brain MRI segmentation and parcellation: Practical approaches and recommendations – insights from the Maastricht study. NeuroImage 237, 118174 (2021).

45. Klapwijk, E. T., van den Bos, W., Tamnes, C. K., Raschle, N. M. & Mills, K. L. Opportunities for increased reproducibility and replicability of developmental neuroimaging. Dev. Cogn. Neurosci. 47, 100902 (2021).

46. Desikan, R. S. et al. An automated labeling system for subdividing the human cerebral cortex on MRI scans into gyral based regions of interest. NeuroImage 31, 968–980 (2006).

47. Cammoun, L. et al. Mapping the human connectome at multiple scales with diffusion spectrum MRI. J. Neurosci. Methods 203, 386–397 (2012).

48. Fortin, J.-P. et al. Harmonization of multi-site diffusion tensor imaging data. NeuroImage 161, 149–170 (2017).

49. Johnson, W. E., Li, C. & Rabinovic, A. Adjusting batch effects in microarray expression data using empirical Bayes methods. Biostatistics 8, 118–127 (2007).

50. Radua, J. et al. Increased power by harmonizing structural MRI site differences with the ComBat batch adjustment method in ENIGMA. NeuroImage 218, 116956 (2020).

51. Hawrylycz, M. J. et al. An anatomically comprehensive atlas of the adult human brain transcriptome. Nature 489, 391–399 (2012).

52. Arnatkeviciute, A., Markello, R. D., Fulcher, B. D., Misic, B. & Fornito, A. Toward Best Practices for Imaging Transcriptomics of the Human Brain. Biol. Psychiatry 93, 391–404 (2023).

53. Quackenbush, J. Microarray data normalization and transformation. Nat. Genet. 32, 496–501 (2002).

54. Hawrylycz, M. et al. Canonical genetic signatures of the adult human brain. Nat. Neurosci. 18, 1832–1844 (2015).

55. Romero-Garcia, R. et al. Structural covariance networks are coupled to expression of genes enriched in supragranular layers of the human cortex. NeuroImage 171, 256–267 (2018).

56. McIntosh, A. R. & Lobaugh, N. J. Partial least squares analysis of neuroimaging data: applications and advances. NeuroImage 23, S250–S263 (2004).

57. Alexander-Bloch, A., Giedd, J. N. & Bullmore, E. Imaging structural co-variance between human brain regions. Nat. Rev. Neurosci. 14, 322–336 (2013).

58. Váša, F. & Mišić, B. Null models in network neuroscience. Nat. Rev. Neurosci. 23, 493– 504 (2022).

59. Efron, B. & Tibshirani, R. Bootstrap Methods for Standard Errors, Confidence Intervals, and Other Measures of Statistical Accuracy. Stat. Sci. 1, 54–75 (1986).

60. Elizarraras, J. M. et al. WebGestalt 2024: faster gene set analysis and new support for metabolomics and multi-omics. Nucleic Acids Res. 52, W415–W421 (2024).

61. Chougar, L. et al. Atrophy in multiple system atrophy relates to mitochondrial and oligodendrocytic processes. Preprint at 10.1101/2025.01.22.25320961 (2025).

62. Eden, E., Navon, R., Steinfeld, I., Lipson, D. & Yakhini, Z. GOrilla: a tool for discovery and visualization of enriched GO terms in ranked gene lists. BMC Bioinformatics 10, 48 (2009).

63. Lonsdale, J. et al. The Genotype-Tissue Expression (GTEx) project. Nat. Genet. 45, 580–585 (2013).

64. Benjamini, Y. & Hochberg, Y. Controlling the False Discovery Rate: A Practical and Powerful Approach to Multiple Testing. J. R. Stat. Soc. Ser. B Stat. Methodol. 57, 289– 300 (1995).

65. Oltra, J. et al. Sex differences in brain atrophy and cognitive impairment in Parkinson’s disease patients with and without probable rapid eye movement sleep behavior disorder. J. Neurol. 269, 1591–1599 (2022).

66. De Micco, R. et al. Sex-related pattern of intrinsic brain connectivity in drug-naïve Parkinson’s disease patients. Mov. Disord. 34, 997–1005 (2019).

67. Bayram, E., Coughlin, D. G., Rajmohan, R. & Litvan, I. Sex differences for clinical correlates of substantia nigra neuron loss in people with Lewy body pathology. Biol. Sex Differ. 15, 8 (2024).

68. Kampmann, M. Molecular and cellular mechanisms of selective vulnerability in neurodegenerative diseases. Nat. Rev. Neurosci. 25, 351–371 (2024).

69. Zarei, M. et al. Cortical thinning is associated with disease stages and dementia in Parkinson’s disease. J. Neurol. Neurosurg. Psychiatry 84, 875–882 (2013).

70. Galli, A. et al. Occipital atrophy signature in prodromal Lewy bodies disease. Alzheimers Dement. Diagn. Assess. Dis. Monit. 15, e12462 (2023).

71. Audet-walsh, É. & Giguére, V. The multiple universes of estrogen-related receptor α and γ in metabolic control and related diseases. Acta Pharmacol. Sin. 36, 51–61 (2015).

72. Sadasivam, N. et al. Exploring the impact of estrogen-related receptor gamma on metabolism and disease. Steroids 211, 109500 (2024).

73. Qian, L. et al. Peroxisome proliferator-activated receptor gamma coactivator-1 (PGC-1) family in physiological and pathophysiological process and diseases. Signal Transduct. Target. Ther. 9, 1–44 (2024).

74. Zheng, B., et al. *PGC-1* α, A Potential Therapeutic Target for Early Intervention in Parkinson’s Disease. Sci. Transl. Med. 2, (2010).

75. Grünewald, A., Kumar, K. R. & Sue, C. M. New insights into the complex role of mitochondria in Parkinson’s disease. Prog. Neurobiol. 177, 73–93 (2019).

76. Fox, S. N. et al. Estrogen-related receptor gamma regulates mitochondrial and synaptic genes and modulates vulnerability to synucleinopathy. Npj Park. Dis. 8, 1–19 (2022).

77. Ciron, C. et al. PGC-1α activity in nigral dopamine neurons determines vulnerability to α-synuclein. Acta Neuropathol. Commun. 3, 16 (2015).

78. McMeekin, L. J. et al. Estrogen-related Receptor Alpha (ERRα) is Required for PGC-1α-dependent Gene Expression in the Mouse Brain. Neuroscience 479, 70–90 (2021).

79. Strosznajder, A. K., Wójtowicz, S., Jeżyna, M. J., Sun, G. Y. & Strosznajder, J. B. Recent Insights on the Role of PPAR-β/δ in Neuroinflammation and Neurodegeneration, and Its Potential Target for Therapy. NeuroMolecular Med. 23, 86–98 (2021).

80. Altinoz, M. A. et al. PPARδ and its ligand erucic acid may act anti-tumoral, neuroprotective, and myelin protective in neuroblastoma, glioblastoma, and Parkinson’s disease. Mol. Aspects Med. 78, 100871 (2021).

81. Chen, L. et al. PPARß/δ agonist alleviates NLRP3 inflammasome-mediated neuroinflammation in the MPTP mouse model of Parkinson’s disease. Behav. Brain Res. 356, 483–489 (2019).

82. Dickey, A. S. et al. PPAR-δ is repressed in Huntington’s disease, is required for normal neuronal function and can be targeted therapeutically. Nat. Med. 22, 37–45 (2016).

83. Kalinin, S., Richardson, J. C. & Feinstein, D. L. A PPARdelta Agonist Reduces Amyloid Burden and Brain Inflammation in a Transgenic Mouse Model of Alzheimer’s Disease. Curr. Alzheimer Res. 6, 431–437 (2009).

