## Supplementary materials for "Estrogen-related receptor genes underlie sex differences in cortical atrophy associated with isolated REM sleep behavior disorder"

### Supplementary material

**Supplementary Material 1. MRI acquisition parameters for each site.**

T1-weighted MRI scans were acquired across sites using 3T MRI scanners with varying acquisition protocols. In the Montreal cohort, scans were obtained using a 3T Siemens TIM Trio scanner with 12-channel head coil and an MPRAGE sequence (TR = 2300 ms, TE = 2.91 ms, flip angle = 9°, isotropic voxel size = 1 mm) or a 3T Siemens PRISMA scanner with a 32-channel head coil and an MPRAGE sequence (TR = 2300 ms, TE = 2.98 ms, flip angle = 9°, isotropic voxel size = 1 mm). The Oxford cohort used a 3T Siemens Trio MRI Scanner with a 12-channel head coil and an MPRAGE sequence (TR = 2040 ms, TE = 4.7 ms, flip angle = 8°, isotropic voxel size = 1 mm). The Prague cohort used a 3T Siemens Skyra with a 32-channel head coil and an MPRAGE sequence (TR = 2200 ms, TE = 2.4 ms, flip angle = 8°, isotropic voxel size = 1 mm). The Paris cohort used a 3T Siemens TIM Trio scanner with a 12-channel head coil and an MPRAGE sequence (TR = 2300 ms, TE = 4.18 ms, flip angle = 9°, isotropic voxel size =1 mm) or a 3T PRISMA Fit scanner with a 64-channel head coil and an MP2RAGE sequence (TR = 5000 ms, TE = 2.98 ms, flip angles = 4° and 5°, GRAPPA = 3, isotropic voxel size = 1 mm). The Sydney cohort used a GE Discovery MR750 3T scanner with an 8-channel head coil and a BRAVO sequence (TR = 5800 ms, TE = 2.6 ms, flip angle = 12°, isotropic voxel size = 1 mm). The Aarhus cohort used a 3T Siemens MAGNETOM Skyra scanner with a 32-channel head coil and an MPRAGE sequence (TR = 2420 ms, TE = 3.7 ms, flip angle = 9°, isotropic voxel size = 1 mm). The Genoa cohort used a 3T Siemens PRISMA Scanner with a 64-channel head coil and an MPRAGE sequence (TR = 2300 ms, TE = 2.98 ms, flip angle = 9°, isotropic voxel size = 1 mm). The acquisition parameters for the PPMI study have been described elsewhere.

**Supplementary Material 2. Detailed surface-based MRI processing.**

To assess the presence of sex-related effects on cortical morphology in iRBD patients, T1-weighted MRI scans were processed to generate whole-brain vertex-based cortical thickness maps for each individual, following previously established methods.[1,2] Cortical reconstruction was performed using the FreeSurfer image analysis suite, with technical details of the procedures described elsewhere.[1,3,4]

Briefly, the processing pipeline included motion correction, [3] removal of non-brain tissue using a hybrid watershed/surface deformation procedure, [4] automated Talairach transformation, segmentation of subcortical white and deep grey matter structures, [5,6] intensity normalization, [7] tessellation of the grey-white matter boundary, automated topology correction,[8,9] and surface deformation guided by intensity gradients to delineate the grey-white and grey-CSF boundaries. [10–12]

Once cortical models were generated, deformable procedures were applied for data analysis, including surface inflation, [13] registration to a spherical atlas using individual cortical folding patterns to align cortical geometry across subjects [14], parcellation of the cortex based on gyral and sulcal structures, [15,16] and creation of surface-based cortical thickness maps. This method leverages both intensity and continuity information from the entire 3D MRI volume for segmentation and deformation, producing cortical thickness measurements calculated as the closest distance from the grey-white boundary to the grey-CSF boundary at each vertex on the tessellated surface.[12] These maps are derived from spatial intensity gradients across tissue classes rather than absolute signal intensity, enabling the detection of submillimetre differences in cortical thickness between groups. Furthermore, this approach is not limited by the voxel resolution of the original data, allowing for high-precision cortical thickness measurements.

Procedures for cortical thickness measurement have been validated against histological analysis [17] and manual measurements, [18,19] demonstrating strong test-retest reliability across scanner manufacturers and field strengths. [20,21] To ensure data quality, trained raters (S.R. and V.A.) visually inspected surface maps and assigned scores from 1 to 4 based on established guidelines,[22,23] with maps receiving scores above 2 excluded from the analysis.

### Supplementary References


1. Rahayel, S. *et al.* Brain atrophy in prodromal synucleinopathy is shaped by structural connectivity and gene expression. *Brain* **145**, 3162–3178 (2022).
2. Joza, S. *et al.* Distinct brain atrophy progression subtypes underlie phenoconversion in isolated REM sleep behaviour disorder. Preprint at https://doi.org/10.1101/2024.09.05.24313131 (2024).
3. Reuter, M., Rosas, H. D. & Fischl, B. Highly accurate inverse consistent registration: A robust approach. *NeuroImage* **53**, 1181–1196 (2010).
4. Ségonne, F. *et al.* A hybrid approach to the skull stripping problem in MRI. *NeuroImage* **22**, 1060–1075 (2004).
5. Fischl, B. *et al.* Whole Brain Segmentation. *Neuron* **33**, 341–355 (2002).
6. Fischl, B. *et al.* Sequence-independent segmentation of magnetic resonance images. *NeuroImage* **23**, S69–S84 (2004).
7. Sled, J. G., Zijdenbos, A. P. & Evans, A. C. A nonparametric method for automatic correction of intensity nonuniformity in MRI data. *IEEE Trans. Med. Imaging* **17**, 87–97 (1998).
8. Fischl, B., Liu, A. & Dale, A. M. Automated manifold surgery: constructing geometrically accurate and topologically correct models of the human cerebral cortex. *IEEE Trans. Med. Imaging* **20**, 70–80 (2001).
9. Segonne, F., Pacheco, J. & Fischl, B. Geometrically Accurate Topology-Correction of Cortical Surfaces Using Nonseparating Loops. *IEEE Trans. Med. Imaging* **26**, 518–529 (2007).
10. Dale, A. M. & Sereno, M. I. Improved Localizadon of Cortical Activity by Combining EEG and MEG with MRI Cortical Surface Reconstruction: A Linear Approach. *J. Cogn. Neurosci.* **5**, 162–176 (1993).

11. Dale, A. M., Fischl, B. & Sereno, M. I. Cortical Surface-Based Analysis: I. Segmentation and Surface Reconstruction. *NeuroImage* **9**, 179–194 (1999).
12. Fischl, B. & Dale, A. M. Measuring the thickness of the human cerebral cortex from magnetic resonance images. *Proc. Natl. Acad. Sci.* **97**, 11050–11055 (2000).
13. Fischl, B., Sereno, M. I. & Dale, A. M. Cortical Surface-Based Analysis. *NeuroImage* **9**, 195–207 (1999).
14. Fischl, B., Sereno, M. I., Tootell, R. B. H. & Dale, A. M. High-resolution intersubject averaging and a coordinate system for the cortical surface. *Hum. Brain Mapp.* **8**, 272–284 (1999).
15. Desikan, R. S. *et al.* An automated labeling system for subdividing the human cerebral cortex on MRI scans into gyral based regions of interest. *NeuroImage* **31**, 968–980 (2006).
16. Fischl, B. Automatically Parcellating the Human Cerebral Cortex. *Cereb. Cortex* **14**, 11–22 (2004).
17. Rosas, H. D. *et al.* Regional and progressive thinning of the cortical ribbon in Huntington's disease. *Neurology* **58**, 695–701 (2002).
18. Kuperberg, G. R. *et al.* Regionally Localized Thinning of the Cerebral Cortex in Schizophrenia. *Arch. Gen. Psychiatry* **60**, 878 (2003).
19. Salat, D. H. Thinning of the Cerebral Cortex in Aging. *Cereb. Cortex* **14**, 721–730 (2004).
20. Han, X. *et al.* Reliability of MRI-derived measurements of human cerebral cortical thickness: The effects of field strength, scanner upgrade and manufacturer. *NeuroImage* **32**, 180–194 (2006).
21. Reuter, M., Schmansky, N. J., Rosas, H. D. & Fischl, B. Within-subject template estimation for unbiased longitudinal image analysis. *NeuroImage* **61**, 1402–1418 (2012).

22. Monereo-Sánchez, J. *et al.* Quality control strategies for brain MRI segmentation and parcellation: Practical approaches and recommendations - insights from the Maastricht study. *NeuroImage* **237**, 118174 (2021).

23. Klapwijk, E. T., van den Bos, W., Tamnes, C. K., Raschle, N. M. & Mills, K. L. Opportunities for increased reproducibility and replicability of developmental neuroimaging. *Dev. Cogn. Neurosci.* **47**, 100902 (2021).

### SUPPLEMENTARY TABLES

**Table S1.** Sex-by-group interaction on cortical thickness.

| Cluster peak location | iRBD | | P-value[a] | Controls | | P-value[b] | Total | | P-value[c] |
|---|---|---|---|---|---|---|---|---|---|
| | Females | Males | | Females | Males | | Controls | iRBD | |
| Left posterior cingulate cortex | 2.49 ±0.11 | 2.43 ±0.11 | **<0.001** | 2.49 ±0.11 | 2.48 ±0.12 | 0.28 | 2.48 ±0.11 | 2.44 ±0.11 | **<0.001** |
| Left superior parietal cortex | 2.18 ±0.11 | 2.11 ±0.12 | **<0.001** | 2.20 ±0.12 | 2.16 ±0.13 | **0.001** | 2.17 ±0.13 | 2.12 ±0.12 | **<0.001** |
| Right paracentral cortex | 2.45 ±0.11 | 2.39 ±0.12 | **<0.001** | 2.47 ±0.12 | 2.45 ±0.13 | 0.16 | 2.46 ±0.13 | 2.40 ±0.12 | **<0.001** |

Data are presented as mean ± SD.
[a] Independent two-sample t-test comparing female and male iRBD patients.
[b] Independent two-sample t-test comparing female and male controls.
[c] Independent two-sample t-test comparing iRBD patients and controls.

iRBD = isolated REM sleep behavior disorder; SD = standard deviation

**Table S2**. Sex effect on cortical thickness in iRBD.

| Cluster peak location | iRBD | | P-value[a] |
|---|---|---|---|
| | Females | Males | |
| Left postcentral cortex | 2.36 ±0.10 | 2.29 ±0.10 | **<0.001** |
| Left inferior parietal cortex | 2.33 ±0.12 | 2.27 ±0.11 | **<0.001** |
| Right caudal middle frontal cortex | 2.42 ±0.11 | 2.36 ±0.13 | **<0.001** |
| Right inferior parietal cortex | 2.19 ±0.14 | 2.12 ±0.14 | **<0.001** |
| Right rostral middle frontal cortex | 2.48 ±0.16 | 2.41 ±0.14 | **<0.001** |
| Right paracentral cortex | 2.40 ±0.16 | 2.34 ±0.16 | **0.007** |

Data are presented as mean ± SD.
[a] Independent two-sample t-test comparing female and male iRBD patients.

iRBD = isolated REM sleep behavior disorder; SD = standard deviation.

**Table S3.** Vertex-based cortical surface area and volume analyses of sex effects

| Cortical measure | Cluster peak location[a] | Cluster size, $mm^2$ | Number of vertices | Talairach coordinates | | | $-\log_{10}$ *P*-value |
|---|---|---|---|---|---|---|---|
| | | | | *x* | *y* | *z* | |
| **Sex-by-group interaction** | | | | | | | |
| Cortical surface area | Right superior frontal cortex | 5353 | 10586 | 24.9 | 6.7 | 46.4 | 5.93 |
| Cortical volume | Right superior frontal cortex | 5012 | 9527 | 21.8 | 13.4 | 48.9 | 6.42 |
| **Sex effect in iRBD** | | | | | | | |
| Cortical surface area | Right superior frontal cortex | 5639 | 11231 | 24.7 | 7.2 | 46.3 | -6.24 |
| Cortical volume | Right superior frontal cortex | 5152 | 9676 | 22.3 | 15.3 | 48.9 | −6.54 |

Clusters were significant after Monte-Carlo simulation, with cluster- and vertex-level P-values set at $P < 0.05$.
[a] Only the region of the peak vertex is indicated (see Figure S2 for mapping).

iRBD = isolated REM sleep behavior disorder.

**Table S4.** Gene association within olfactory receptor activity for LV3.

| Gene | Bootstrap ratio | Gene symbol | Gene name |
|---|---|---|---|
| OR2L13 | 3.15 | OR2L13 | olfactory receptor family 2 subfamily L member 13 |
| OR1E1 | 2.74 | OR1E1 | olfactory receptor family 1 subfamily E member 1 |
| OR10A2 | 2.48 | OR10A2 | olfactory receptor family 10 subfamily A member 2 |
| OR2H1 | 2.24 | OR2H1 | olfactory receptor family 2 subfamily H member 1 |
| OR2AK2 | 2.22 | OR2AK2 | olfactory receptor family 2 subfamily AK member 2 |
| OR2L8 | 2.18 | OR2L8 | olfactory receptor family 2 subfamily L member 8 |
| OR2L3 | 2.17 | OR2L3 | olfactory receptor family 2 subfamily L member 3 |
| OR10H2 | 2.03 | OR10H2 | olfactory receptor family 10 subfamily H member 2 |
| OR1J4 | 1.75 | OR1J4 | olfactory receptor family 1 subfamily J member 4 |
| OR8B12 | 1.70 | OR8B12 | olfactory receptor family 8 subfamily B member 12 |
| OR5L2 | 1.61 | OR5L2 | olfactory receptor family 5 subfamily L member 2 |
| OR4D6 | 1.54 | OR4D6 | olfactory receptor family 4 subfamily D member 6 |
| OR3A1 | 1.50 | OR3A1 | olfactory receptor family 3 subfamily A member 1 |
| OR2AG2 | 1.44 | OR2AG2 | olfactory receptor family 2 subfamily AG member 2 |
| OR52W1 | 1.28 | OR52W1 | olfactory receptor family 52 subfamily W member 1 |
| OR4K2 | 1.24 | OR4K2 | olfactory receptor family 4 subfamily K member 2 |
| OR2H2 | 1.16 | OR2H2 | olfactory receptor family 2 subfamily H member 2 |
| OR6C65 | 1.12 | OR6C65 | olfactory receptor family 6 subfamily C member 65 |
| OR10C1 | 1.12 | OR10C1 | olfactory receptor family 10 subfamily C member 1 |

| | | | |
|---|---|---|---|
| OR5H1 | 1.12 | OR5H1 | olfactory receptor family 5 subfamily H member 1 |
| OR6M1 | 1.11 | OR6M1 | olfactory receptor family 6 subfamily M member 1 |
| OR11A1 | 1.11 | OR11A1 | olfactory receptor family 11 subfamily A member 1 |
| OR4D2 | 1.10 | OR4D2 | olfactory receptor family 4 subfamily D member 2 |
| OR2T6 | 1.01 | OR2T6 | olfactory receptor family 2 subfamily T member 6 |
| OR2A14 | 1.00 | OR2A14 | olfactory receptor family 2 subfamily A member 14 |
| OR2M7 | 0.92 | OR2M7 | olfactory receptor family 2 subfamily M member 7 |
| OR13A1 | 0.90 | OR13A1 | olfactory receptor family 13 subfamily A member 1 |
| OR3A2 | 0.89 | OR3A2 | olfactory receptor family 3 subfamily A member 2 |
| OR2A2 | 0.87 | OR2A2 | olfactory receptor family 2 subfamily A member 2 |

Only showing the 29 out of 48 gene terms associated with this molecular function.

**Table S5.** Molecular functions associated with positive sex interaction for LV1 in GOrilla.

| GO term | Description | FDR P-value[a] | Enrichment[b] | N | B | n | b |
|---|---|---|---|---|---|---|---|
| GO:0046873 | metal ion transmembrane transporter activity | **0.0005** | 2.09 | 13979 | 318 | 1327 | 63 |
| GO:0003707 | steroid hormone receptor activity | **0.0052** | 14.34 | 13979 | 21 | 325 | 7 |
| GO:0004879 | nuclear receptor activity | **0.0128** | 6.14 | 13979 | 39 | 642 | 11 |
| GO:0098531 | transcription factor activity, direct ligand regulated sequence-specific DNA binding | **0.0096** | 6.14 | 13979 | 39 | 642 | 11 |
| GO:0008324 | cation transmembrane transporter activity | **0.0126** | 1.74 | 13979 | 449 | 1327 | 74 |
| GO:0022890 | inorganic cation transmembrane transporter activity | **0.0191** | 1.75 | 13979 | 410 | 1327 | 68 |
| GO:0005249 | voltage-gated potassium channel activity | **0.0206** | 3.26 | 13979 | 71 | 1148 | 19 |
| GO:0001227 | DNA-binding transcription repressor activity, RNA polymerase II-specific | **0.0345** | 1.93 | 13979 | 250 | 1333 | 46 |
| GO:1905056 | calcium-transporting ATPase activity involved in regulation of presynaptic cytosolic calcium ion concentration | **0.0323** | 194.15 | 13979 | 3 | 48 | 2 |
| GO:0005251 | delayed rectifier potassium channel activity | **0.0497** | 10.51 | 13979 | 23 | 347 | 6 |
| GO:0015077 | monovalent inorganic cation transmembrane transporter activity | 0.0511 | 1.89 | 13979 | 260 | 1281 | 45 |
| GO:0015079 | potassium ion transmembrane transporter activity | 0.0551 | 2.6 | 13979 | 122 | 1057 | 24 |

| | | | | | | | |
|---|---|---|---|---|---|---|---|
| GO:0005244 | voltage-gated ion channel activity | 0.0683 | 3.18 | 13979 | 153 | 460 | 16 |
| GO:0022832 | voltage-gated channel activity | 0.0634 | 3.18 | 13979 | 153 | 460 | 16 |
| GO:0005496 | steroid binding | 0.0769 | 6.63 | 13979 | 74 | 228 | 8 |
| GO:0022843 | voltage-gated cation channel activity | 0.0898 | 2.6 | 13979 | 112 | 1057 | 22 |
| GO:0015318 | inorganic molecular entity transmembrane transporter activity | 0.1030 | 1.53 | 13979 | 578 | 1327 | 84 |
| GO:0005388 | calcium-transporting ATPase activity | 0.2040 | 83.21 | 13979 | 7 | 48 | 2 |
| GO:0047389 | glycerophosphocholine phosphodiesterase activity | 0.1950 | 2329.83 | 13979 | 2 | 3 | 1 |
| GO:2001070 | starch binding | 0.1850 | 2329.83 | 13979 | 2 | 3 | 1 |
| GO:0052798 | beta-galactoside alpha-2,3-sialyltransferase activity | 0.1780 | 1164.92 | 13979 | 1 | 12 | 1 |
| GO:0005261 | cation channel activity | 0.1890 | 1.79 | 13979 | 235 | 1327 | 40 |

Bold values represent significantly enriched gene terms.
[a] Correction of the p-value for multiple testing using the Benjamini and Hochberg (1995) method.
[b] Enrichment scores calculated as (b/n) / (B/N).

GO = Gene Ontology; FDR = False Discovery Rate; N = total number of genes; B = total number of genes associated with a specific GO term; n = number of genes in the top of ranked gene list; b = number of genes in the intersection.

**Table S6.** Gene association within steroid hormone receptor function for LV1 in GOrilla.

| User ID | Gene symbol | Gene name |
|---|---|---|
| NR3C2 | NR3C2 | Nuclear receptor subfamily 3, group c, member 2 |
| NR2C1 | NR2C1 | Nuclear receptor subfamily 2, group c, member 1 |
| ESRRA | ESRRA | Oestrogen-related receptor alpha |
| ESRRG | ESRRG | Oestrogen-related receptor gamma |
| PPARD | PPARD | Peroxisome proliferator-activated receptor delta |
| NR3C1 | NR3C1 | Nuclear receptor subfamily 3, group c, member 1 (glucocorticoid receptor) |
| NKX3-1 | NKX3-1 | Nk3 homeobox 1 |

**Table S7.** Gene association within nuclear steroid receptor function for LV1 in GOrilla.

| Gene ID | Gene symbol | Gene name |
|---|---|---|
| NR3C2 | NR3C2 | Nuclear receptor subfamily 3, group c, member 2 |
| RORA | RORA | Rar-related orphan receptor a |
| NR2C1 | NR2C1 | Nuclear receptor subfamily 2, group c, member 1 |
| ESRRA | ESRRA | Oestrogen-related receptor alpha |
| RORB | RORB | Rar-related orphan receptor b |
| ESRRG | ESRRG | Oestrogen-related receptor gamma |
| NR1D2 | NR1D2 | Nuclear receptor subfamily 1, group d, member 2 |
| NR1D1 | NR1D1 | Nuclear receptor subfamily 1, group d, member 1 |
| PPARD | PPARD | Peroxisome proliferator-activated receptor delta |
| NR3C1 | NR3C1 | Nuclear receptor subfamily 3, group c, member 1 (glucocorticoid receptor) |

### SUPPLEMENTARY FIGURES

**Figure S1.** Sex effect on cortical thickness in iRBD.

(a) Clusters showing significant sex effect on cortical thickness in iRBD. The colour bar indicates the statistical significance on a logarithmic scale of *P*-values (−log10), with positive values showing significant decreases in iRBD males compared to iRBD females. (b) Average cortical thickness (in mm) in iRBD males and females in significant clusters, showing significant reduction in cortical thickness in iRBD males compared to iRBD females. The P-values indicate significant differences between groups after conducting independent two sample t-tests. See Table S2 for details. iRBD-M = iRBD males; iRBD-F= iRBD females.

**Figure S2.** Vertex-wise analyses on cortical surface area and cortical volume.

(a) Clusters showing significant sex-group interaction on cortical surface area and cortical volume. The colour bar indicates the statistical significance on a logarithmic scale of *P*-values (−log10), with positive values showing significant decreases in iRBD males compared to females and controls. (b) Clusters showing significant sex effect on cortical surface area and cortical volume in the iRBD group only. The colour bar indicates the statistical significance on a logarithmic scale of *P*-values (−log10), with negative values showing significant decreases in iRBD males compared to iRBD females (iRBD males < iRBD females)

**Figure S3.** Top 24 tissue expression of *PPARD*.

Tissue expression values of *PPARD*, showing a more ubiquitous distribution across tissue types (lilac), and not brain specific (yellow) pattern of expression. Box plots are shown as median, 25$^{th}$ and 75$^{th}$ percentiles. Points appearing outside of the plots corresponds to outliers above or below 1.5 times the interquartile range. TPM = transcripts per million.
