## Supplementary figures and images for "Estrogen-related receptor genes underlie sex differences in cortical atrophy associated with isolated REM sleep behavior disorder"

### Supplementary figure 1

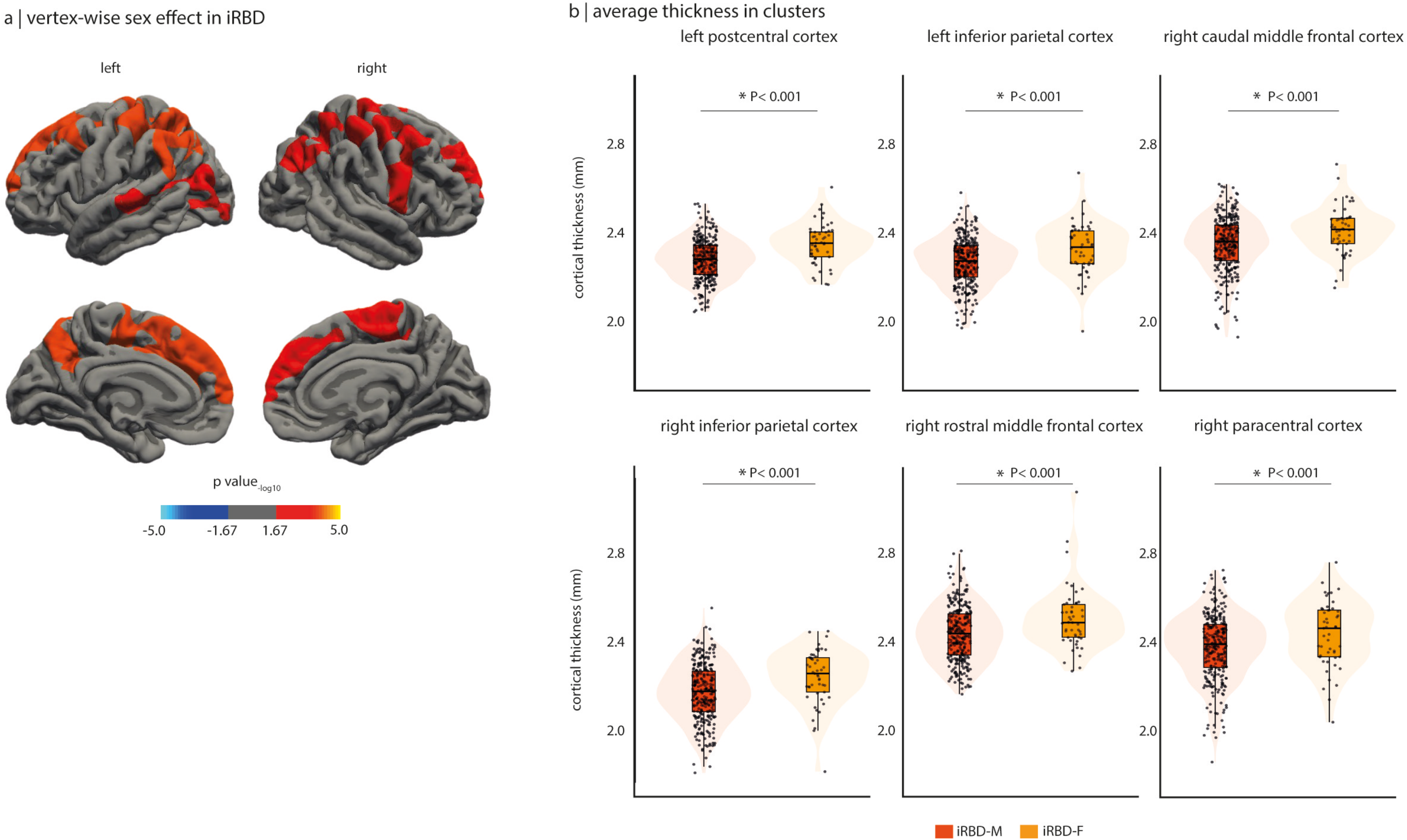

### Supplementary figure 2

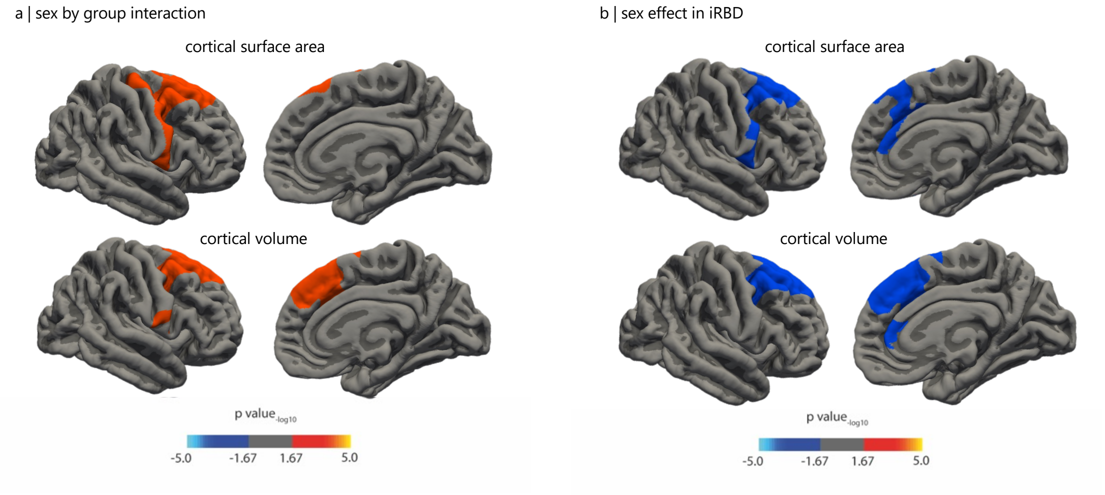

### Supplementary figure 3

a | bulk tissue gene expression for *PPARD*

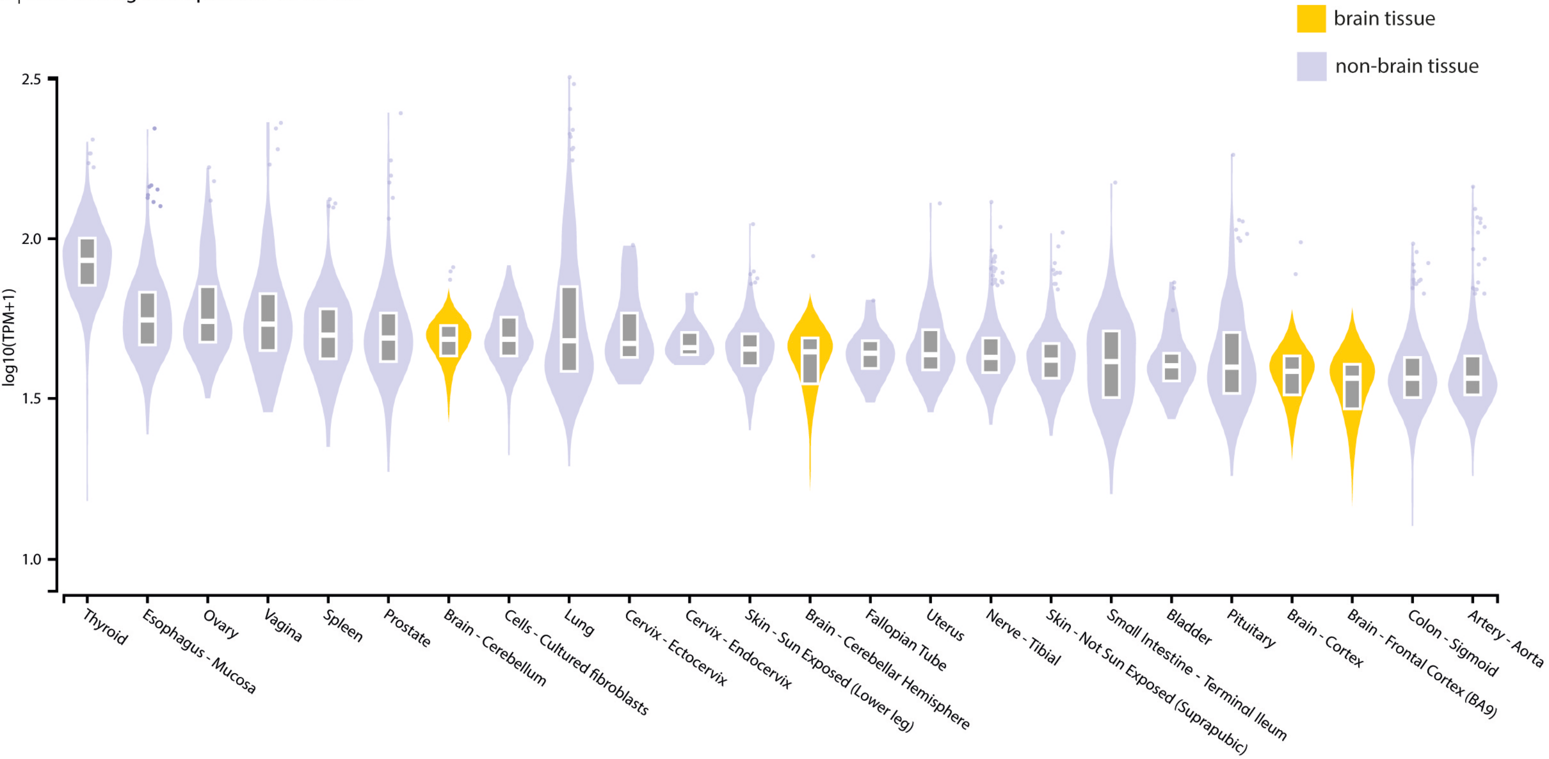
